# Sex-specific metabolic pathways associate with Alzheimer’s Disease (AD) endophenotypes in the European Medical Information Framework for AD Multimodal Biomarker Discovery cohort

**DOI:** 10.1101/2021.04.06.21254535

**Authors:** Jin Xu, Rebecca Green, Min Kim, Jodie Lord, Amera Ebshiana, Sarah Westwood, Alison L Baird, Alejo J Nevado-Holgado, Liu Shi, Abdul Hye, Stuart G. Snowden, Isabelle Bos, Stephanie J. B. Vos, Rik Vandenberghe, Charlotte E. Teunissen, Mara Ten Kate, Philip Scheltens, Silvy Gabel, Karen Meersmans, Olivier Blin, Jill Richardson, Ellen Elisa De Roeck, Sebastiaan Engelborghs, Kristel Sleegers, Régis Bordet, Lorena Rami, Petronella Kettunen, Magda Tsolaki, Frans R.J. Verhey, Daniel Alcolea, Alberto Lleó, Gwendoline Peyratout, Mikel Tainta, Peter Johannsen, Yvonne Freund-Levi, Lutz Frölich, Valerija Dobricic, Giovanni B. Frisoni, José Luis Molinuevo, Anders Wallin, Julius Popp, Pablo Martinez-Lage, Lars Bertram, Kaj Blennow, Henrik Zetterberg, Johannes Streffer, Pieter Jelle Visser, Simon Lovestone, Petroula Proitsi, Cristina Legido-Quigley, on behalf of EMIF Consortium

**Affiliations:** Institute of Pharmaceutical Science, King’s College London, London, United Kingdom; Institute of Psychiatry, Psychology and Neuroscience, Maurice Wohl Clinical Neuroscience Institute, King’s College London, London, United Kingdom; NIHR Maudsley Biomedical Research Centre, South London and Maudsley NHS Trust, London, United Kingdom; Steno Diabetes Center, Gentofte, Denmark; Department of Psychiatry, University of Oxford, Oxford, United Kingdom; Department of Neurology, Alzheimer Center Amsterdam, Amsterdam Neuroscience, Vrije Universiteit Amsterdam, Amsterdam, Netherlands; Department of Psychiatry and Neuropsychology, School for Mental Health and Neuroscience, Alzheimer Centrum Limburg, Maastricht University, Maastricht, Netherlands; Department of Radiology and Nuclear Medicine, VU University Medical Center, Amsterdam, Netherlands; Department of Clinical Chemistry, Neurochemistry Laboratory, Amsterdam Neuroscience, Amsterdam University Medical Centers, Vrije Universiteit, Amsterdam, Netherlands; Department of Neurosciences, Laboratory for Cognitive Neurology, Leuven, Belgium; University Hospital Leuven, Leuven, Belgium; Aix-Marseille University-CNRS, Marseille, France; Neurosciences Therapeutic Area, GlaxoSmithKline R&D, Stevenage, United Kingdom; Vrije Universiteit Brussel, Brussels, Belgium; Reference Center for Biological Markers of Dementia (BIODEM), Institute Born-Bunge, University of Antwerp, Antwerp, Belgium; Department of Neurology and Center for Neurosciences (C4N), UZ Brussel and Vrije Universiteit Brussel (VUB), Brussels, Belgium; Institute Born-Bunge, University of Antwerp, Antwerp, Belgium; Complex Genetics of Alzheimer’s Disease Group, VIB Center for Molecular Neurology, VIB, Antwerp, Belgium; Université de Lille, Lille, France; Alzheimer’s Disease and Other Cognitive Disorders Unit, Hospital Clínic of Barcelona, IDIBAPS, Barcelona, Spain; Institute of Neuroscience and Physiology, Sahlgrenska Academy, University of Gothenburg, Gothenburg, Sweden; AHEPA University Hospital, Thessaloniki, Greece; Alzheimer Center Limburg, School for Mental Health and Neuroscience, Maastricht University, Maastricht, Netherlands; Sant Pau Memory Unit, Department of Neurology, Hospital de la Santa Creu i Sant Pau, Barcelona, Spain; Lausanne University Hospital, Lausanne, Switzerland; Fundación CITA-Alzhéimer Fundazioa, San Sebastian, Spain; Danish Dementia Research Centre, Rigshospitalet, Copenhagen, Denmark; Department of Neurobiology, Caring Sciences and Society (NVS), Division of Clinical Geriatrics, Karolinska Institutet, Department of Psychiatry in Region Örebro County and School of Medical Sciences, Faculty of Medicine and Health, Örebro University, Örebro, Sweden; Department of Old Age Psychiatry, Psychology & Neuroscience, King’s College, London, UK; Department of Geriatric Psychiatry, Central Institute of Mental Health, Medical Faculty Mannheim, University of Heidelberg, Mannheim, Germany; Lübeck Interdisciplinary Platform for Genome Analytics, Institutes of Neurogenetics and Cardiogenetics, University of Lübeck, Lübeck, Germany; University of Geneva, Geneva, Switzerland; IRCCS Fatebenefratelli, Brescia, Italy; Barcelona Beta Brain Research Center, Unversitat Pompeu Fabra, Barcelona, Spain; University of Lille, Lille, France; Old Age Psychiatry, Department of Psychiatry, University Hospital Lausanne, Lausanne, Switzerland; Department of Geriatric Psychiatry, University Hospital of Psychiatry Zürich, Zürich, Switzerland; Center for Research and Advanced Therapies, CITA-Alzheimer Foundation, San Sebastian, Spain; University of Oslo, Oslo, Norway; University of Lübeck, Lübeck, Germany; Clinical Neurochemistry Laboratory, Sahlgrenska University Hospital, Mölndal, Sweden; Department of Psychiatry and Neurochemistry, Institute of Neuroscience and Physiology, University of Gothenburg, Mölndal, Sweden; Institute of Neuroscience and Physiology, Department of Psychiatry and Neurochemistry, The Sahlgrenska Academy at University of Gothenburg, Mölndal, Sweden; UK Dementia Research Institute at UCL, London, United Kingdom; Department of Neurodegenerative Disease, UCL Institute of Neurology, London, United Kingdom; Department of Psychiatry and Neuropsychology, School for Mental Health and Neuroscience, Maastricht University, Maastricht, Netherlands; Alzheimer Center Amsterdam, Amsterdam Neuroscience, Vrije Universiteit Amsterdam, Amsterdam UMC, Amsterdam, Netherlands; Janssen-Cilag UK Ltd, Oxford, United Kingdom

**Keywords:** sex, Alzheimer’s disease, metabolomics, metabolic pathway, blood, vanillylmandelate, tryptophan betaine

## Abstract

**BACKGROUND:** Physiological differences between males and females could contribute to the development of AD. Here, we examined metabolic pathways that may lead to precision medicine initiatives.

**METHODS:** We explored whether sex modifies the association of 540 plasma metabolites with AD endophenotypes including diagnosis, CSF biomarkers, brain imaging, and cognition using regression analyses for 695 participants (377 females), followed by sex-specific pathway overrepresentation analyses, *APOE* ε4 stratification and assessment of metabolites’ discriminatory performance in AD.

**RESULTS:** In females with AD, vanillylmandelate (tyrosine pathway) was increased and tryptophan betaine (tryptophan pathway) was decreased. The inclusion of these two metabolites (AUC = 0.83, SE = 0.029) to a baseline model (covariates + CSF biomarkers, AUC = 0.92, SE = 0.019) resulted in a significantly higher AUC of 0.96 (SE = 0.012). Kynurenate was decreased in males with AD (AUC = 0.679, SE = 0.046).

**CONCLUSIONS:** Metabolic sex-specific differences were reported, covering neurotransmission and inflammation pathways with AD endophenotypes. Two metabolites, in pathways related to dopamine and serotonin, associated to females, paving the way to personalised treatment.

## 1 Background

Alzheimer’s disease (AD) is the most common neurodegenerative dementia, making it a major source of global morbidity and mortality. Current clinical diagnosis of AD relies on a battery of cognitive tests combined with structural and functional imaging and cerebrospinal fluid (CSF) biomarkers (1). Studies on mechanisms and biomarkers of AD are attracting considerable interest due to the challenge of diagnosing cognitively normal people during the preclinical phase of the disease.

Biomarkers of AD can be described in the ATN framework, grouped into those of β amyloid (Aβ) deposition, pathological tau, and neurodegeneration (2). This ATN descriptive classification system is independent to clinically defined diagnostic criteria and eases the categorisation of multidomain biomarker findings at the individual level (3). Aβ (measured by CSF Aβ_42/40_ or amyloid PET or CSF Aβ_42_) and tau (evaluated by CSF phosphorylated tau or tau PET) are being refined as the primary mediators of AD-related synaptic loss and eventual neuronal death (4). AD neurodegeneration or neuronal injury can be quantitatively or topographically estimated by CSF t-tau, [^18^F] fluorodeoxyglucose-positron emission tomography (FDG-PET), or structural MRI (5). Although there are challenges of measuring biomarkers for brain diseases in the blood (low concentrations and expression in non-cerebral tissues), blood-based biomarkers are easily accessible, minimally invasive and scalable for large scale use. Recently, success in detecting AD and predicting future disease progression has been achieved particularly for the application of glial fibrillary acidic protein (GFAP), plasma phospho-tau181and phospho-tau217 (6–8).

Additionally to the above-mentioned biomarkers, AD risk factors include age, as well as genetic, environmental and modifiable lifestyle factors (9). Sex-related differences in brain structure and function have also been widely discussed (10,11), and if we know more about whether sex-specific associations exist and the biological underpinnings of these relationships, this may help guide more personalised treatments (12). Studies have suggested that females are at greater risk for developing AD when compared to males. The UK dementia report states that more females have AD and that this trend is likely to remain with a F:M ratio of approximately 2 to 1 in the projected dementia incidence for 2051 (13). Although it has been recently reported that selective survival may contribute to sex/gender differences in dementia incidence, this does not preclude the additional contributions from biological mechanisms (14).

Our previous studies using smaller cohorts have shown that i) plasma metabolites have the potential to match the AUC of well-established AD CSF biomarkers through machine learning approach (sample size N = 357) and ii) primary fatty amides were associated with AD endophenotypes (sample size N = 593) (15,16). In the present study, we performed metabolomics in 102 additional participants to a total of 695 and aimed to explore whether sex modifies the association of plasma metabolites with AD endophenotypes. Linear and logistic regressions were performed to investigate the association of plasma metabolite levels with AD endophenotypes (CSF biomarkers, brain MRI, cognition measures and diagnosis) in the full cohort including a sex interaction term, followed by sex-stratified analyses and pathway analyses. Prediction models based on logistic and linear regression were then built to assess AD biomarker performance in each sex stratum. The study design is illustrated in Supplementary Figure S1.

## 2 Methods

### 2.1 Participants

This study included 695 participants consisting of 377 females and 318 males from the European Medical Information Framework for Alzheimer’s Disease Multimodal Biomarker Discovery (EMIF-AD MBD) (17). There were 283 control (CTL) participants, 275 participants with mild cognitive impairment (MCI) and 137 AD dementia patients. Participants were included from three multicenter studies: EDAR (n = 84) (18), PharmaCog (n = 40) (19), DESCRIPA (n = 16) (20), and eight single center studies: Amsterdam (n = 147) (21), Antwerp (n =133) (22), IDIBAPS (n = 93) (23), Leuven (n = 53) (24), San Sebastian GAP (n = 40) (25), Barcelona-Sant Pau (n = 35) (26), Gothenburg (n = 34) (27), Lausanne (n = 20) (28).

### 2.2 Clinical and cognitive data

Neuropsychological tests measuring five different cognitive domains were available. Briefly, these were memory (delayed and immediate), language, attention, executive function, and visuo-construction (Table 1). Hippocampal volume (left, right and sum, adjusted for intracranial volume ICV) and cortical thickness (average across the whole brain and in AD signature regions) were also available (Table 1). The definition of CTL was a normal performance on neuropsychological assessment (within 1.5 SD of the average for age, gender and education). MCI was defined as having cognitive complaints and performance below 1.5 SD of the average on at least one neuropsychological test but no dementia (29). AD-type dementia diagnosis was made based on a clinical diagnosis, using the National Institute of Neurological and Communicative Disorders and Stroke – Alzheimer’s Disease and Related Disorders Association criteria (30). Details on the neuropsychological test, amyloid (CSF Aβ_42/40_ ratio of the central analyses or the local CSF Aβ_42_ value or the standardized uptake value ratio (SUVR) on an amyloid-PET scan) and tau level measurements, magnetic resonance imaging (MRI) and genetic analyses can be found in Bos *et al*. (17).

**Table 1.** Sample demographics.

|  | Sample size | CTL | MCI | AD | P- value* |
| --- | --- | --- | --- | --- | --- |
| Age | 695 | 65 (7.9) | 70 (8.1) | 70 (8.49) | 6.52e-14 |
| Sex (f/m) | 377/318 | 155/128 | 141/134 | 81/56 | 3.13e-01 |
| *APOE* $\epsilon 4$ (+/-) | 348/347 | 111/172 | 153/122 | 84/53 | 7.30e-06 |
| Aβ z-score | 693 | -0.22 (1.11) | -0.82 (0.99) | -1.22 (0.64) | <2.00e-16 |
| p-tau z-score | 640 | -0.00087 (0.99) | -0.94 (1.38) | -1.31 (1.66) | <2.00e-16 |
| t-tau z-score | 637 | 0.032 (0.84) | -0.98 (1.25) | -1.65 (1.60) | <2.00e-16 |
| Attention z-score | 644 | 0.21 (1.13) | -0.91 (1.63) | -1.92 (1.99) | <2.00e-16 |
| Executive z-score | 526 | 0.16 (1.16) | -0.79 (2.04) | -2.26 (2.55) | <2.00e-16 |
| Language z-score | 674 | -0.18 (0.99) | -1.0 (1.25) | -2.08 (1.27) | <2.00e-16 |
| Memory delayed z-score | 551 | -0.037 (1.15) | -1.52 (1.40) | -2.40 (1.070) | <2.00e-16 |
| Memory immediate z-score | 637 | -0.50 (1.77) | -1.57 (1.39) | -2.34 (1.24) | <2.00e-16 |
| Visuo-construction z-score | 436 | -0.20 (1.34) | -0.14 (1.47) | -1.36 (1.98) | 1.93e-08 |
| Hippocampal left | 455 | 3761.21 (453.57) | 3272.52 (634.570) | 3017.90 (487.63) | <2.00e-16 |
| Hippocampal right | 455 | 3878.12 (436.76) | 3388.14 (628.15) | 3146.17 (500.52) | <2.00e-16 |
| Hippocampal sum | 455 | 7639.32 (857.95) | 6660.69 (1210.53) | 6182.10 (913.81) | <2.00e-16 |
| Cortical thickness in whole brain | 420 | 2.30 (0.12) | 2.30 (0.11) | 2.28 (0.11) | 5.47e-01 |
| Cortical thickness in AD regions | 420 | 2.66 (0.16) | 2.63 (0.15) | 2.58 (0.17) | 1.95e-04 |
| Taking AChEI, yes/no | 76/149 | 1/40 | 50/78 | 25/31 | 1.3e-05 |
| Taking other AD medications, yes/no | 23/201 | 0/41 | 16/112 | 7/48 | 1.55e-01 |

Results are mean (standard deviation) for continuous variables.
Aβ , amyloid beta; AChEI, acetylcholine esterase inhibitor; AD, Alzheimer's disease; APOE $\epsilon 4$ , apolipoproteinE $\epsilon 4$ ; MCI, mild cognitive impairment; MMSE, Mini-Mental State Examination; CTL, control; ROD, rate of cognitive decline.
\*Differences in the means/frequencies of clinical/demographic variables were tested using ANOVA or $\chi^2$ test.

### 2.3 Metabolomics data

Relative levels of 665 plasma metabolites were measured in fasted blood samples by *Metabolon, Inc*. Metabolomics experiments were performed with an additional 102 participants, and details of the analytical method for all (N=695) can be found in Kim *et al*. (16). After removing metabolites with more than 20% missing values, 540 metabolites with known identity were included for further analyses. Log10 transformation and scaling (mean of zero and standard deviation of one) were applied to metabolomics data after missing values were imputed with the k-nearest neighbour algorithm (k=10, “impute” package for R). Single outlying values beyond three standard deviations were further removed.

### 2.4 Statistical analyses

Prior to statistical analyses, baseline characteristics were compared among the three diagnostic groups using the ANOVA for continuous variables and Chi-square test for categorical variables (Table 1). The 15 AD endophenotypes were classified into four collections: CSF biomarkers (three continuous variables; Aβ, p-tau, t-tau), brain MRI measurements (five continuous variables; hippocampal left, right and sum volume, average cortical thickness in whole brain and AD regions), cognition measures (six continuous variables; attention, executive, language, memory delay, memory immediate, visuoconstruction) and diagnosis (one binary variable; AD versus CTL, MCI excluded).

Regression analyses evaluated the association between metabolites (predictors) and AD endophenotypes (outcomes). Linear regression was used for continuous AD variables and logistic regression was used for diagnosis. An interaction term between sex and metabolites was included in each model to test whether sex modified the association of metabolites with AD endophenotypes. All models were further sex-stratified when there was evidence of significant sex-metabolite interaction (P<0.05). Each regression model was adjusted for age at sampling, years of education, batch and presence of the *APOE* ε*4* allele (ε*4* carrier versus no ε*4* carrier). The analyses of *APOE* ε*4* (+/-) -by-metabolite interaction effects were also conducted using the same approach described above where sex was included as a covariate instead (Supplementary methods and results). To account for the number of tests performed and the correlations between metabolites, a Bonferroni-adjusted significance threshold (P <8.17e-04) was set as 0.05 divided by the number of independent metabolites (n = 61) from 230 metabolites used in sex-stratified analyses (“independent tests” package in Python https://github.com/hagax8/independent_tests). Exploratory pathway enrichment analyses were additionally performed (Supplementary methods and results).

Individual metabolites showing association with diagnosis (P<8.17e-04) were investigated as sex-specific biomarkers using prediction models (bootstrapped, n=1000, “fbroc” package in R). To evaluate the prediction performance gained by adding sex-specific biomarkers to baseline AD predictors, likelihood ratio test (“lmtest” package in R) was employed followed by net reclassification index (NRI) analysis (“nricens” package in R). The baseline model was built including both CSF biomarkers and covariates. Sex-specific markers were then added, and their performance was evaluated. The prevalence of AD in the dataset (excluding MCI) was utilised as the event rate and participants were categorised into two groups: below and above the event rate. The NRI categorized at the event rate, denoted NRI(p), is a robust measure to model miscalibration (31). We calculated both event NRI and non-event NRI and took the sum as the final NRI(p) (32). It has been reported that adding a variable/variables that has/have a moderate or large effect size (Cohen’s D equal to 0.5 or 0.8, respectively) can yield NRI(p) values between 0.018 and 0.197, depending on the discrimination of the initial model (AUC) (33). To provide insight into the links between diagnosis and other AD endophenotypes, metabolites that were nominally associated with diagnosis and at least one other AD variable were studied further. All statistical analyses were performed using R Studio (version 1.3.1056) unless otherwise stated.

## 3 Results

### 3.1 Demographics

The characteristics of the study participants are presented in Table 1. There were no differences in sex distribution among the three diagnostic groups. AD and MCI participants were older compared to CTL participants. There were more *APOE ε4* carriers in the AD group compared to the other two groups (*P* < 0.01). AD participants had lower CSF Aβ, p-tau, and t-tau z-score levels (*P* < 0.01) compared to the other two groups, which means lower CSF Aβ_42_ and CSF Aβ_42/40_ ratio. AD participants also had smaller hippocampal volumes (left, right, and sum) and on average smaller cortical thickness in AD signature regions (all, *P* < 0.01). No significant differences were observed for average cortical thickness across the whole brain between the three diagnostic groups (*P* ≥ 0.05).

### 3.2 Sex-specific association of blood metabolites with AD endophenotypes

To identify whether sex modified the association between blood metabolites and the AD endophenotypes included in this study, we built multivariable linear and logistic regression models between each of the 540 metabolite and each of the 15 AD endophenotypes from four collections (CSF biomarkers, brain MRI measurements, cognition measures and diagnosis) by including an interaction term between sex and each metabolite. We found that sex modified the association of 230 metabolites with AD endophenotypes (Supplementary Table S1). Of these, 38 metabolites showed sex-interactions with CSF biomarkers, 152 with cognition measurements, 79 with brain MRI variables, and 30 with diagnosis (AD/CTL).

Metabolites that showed an interaction with sex at P<0.05 were further investigated for associations with AD endophenotypes in sex-stratified regression analyses. Specifically, only metabolites that showed an interaction with sex in the full dataset, and that associated with the same endophenotype in our sex-stratified analyses, were listed. Stratifying participants by sex revealed that two metabolites, vanillylmandelate (VMA) and tryptophan betaine (Figure 2A & 2B), were associated with AD in females after Bonferroni correction (P < 8.17e-04) (β = 0.77, 95% CI = 0.40 to 1.18, P = 1.14e-04 and β = -0.73, 95% CI = -1.12 to -0.36, P = 1.48e-04, respectively), and 116 of 230 metabolites at the nominal significance level. In parallel, one metabolite, kynurenate, was associated with AD in males after Bonferroni correction (P < 8.17e-04) (β = -1.04, 95% CI = -1.58 to -0.54, P = 7.63e-05) (Table 2) and 100 of 230 metabolites showed nominally significant associations in males (Supplementary Table S1).

**Table 2.**
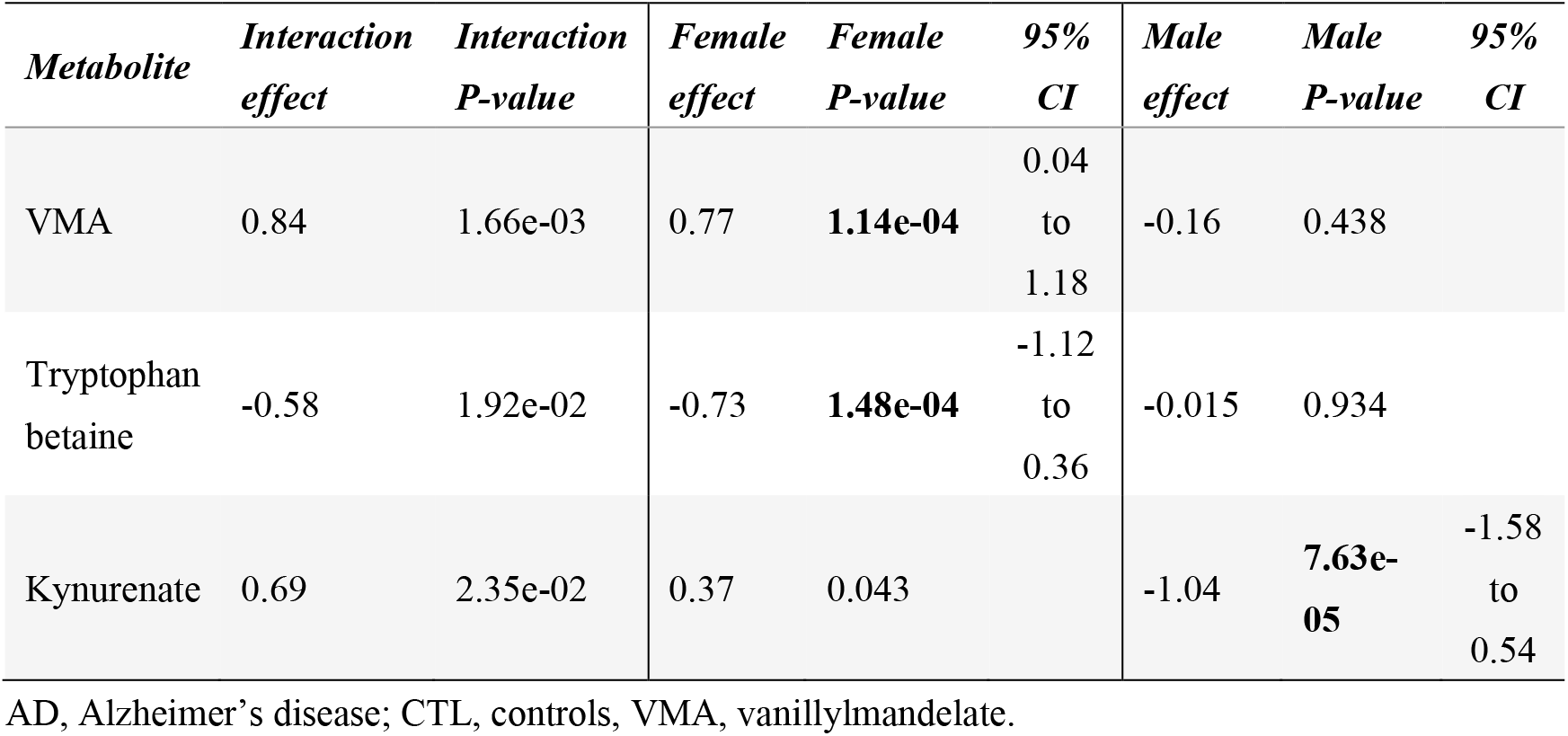
Association results for metabolites in relation to diagnosis (AD/CTL) in the full cohort and after stratification by sex.

| *Metabolite* | *Interaction effect* | *Interaction P-value* | *Female effect* | *Female P-value* | *95% CI* | *Male effect* | *Male P-value* | *95% CI* |
| --- | --- | --- | --- | --- | --- | --- | --- | --- |
| VMA | 0.84 | 1.66e-03 | 0.77 | **1.14e-04** | 0.04 to 1.18 | -0.16 | 0.438 |  |
| Tryptophan betaine | -0.58 | 1.92e-02 | -0.73 | **1.48e-04** | -1.12 to 0.36 | -0.015 | 0.934 |  |
| Kynurenate | 0.69 | 2.35e-02 | 0.37 | 0.043 |  | -1.04 | **7.63e-05** | -1.58 to 0.54 |

AD, Alzheimer's disease; CTL, controls, VMA, vanillylmandelate.

In addition to the single metabolite analysis, pathway-based exploratory analysis was performed to provide more insights into these metabolites in AD at a systems level (Supplementary methods and results). We also investigated metabolites that showed sex-specific associations with diagnosis and at least one other AD phenotype (P < 0.05). As shown in Figure 1, five metabolites showed associations only in females. These included isovalerylcarnitine from ‘leucine, isoleucine and valine metabolism’ sub-pathway, carotene diol (1) and carotene diol (2) from ‘vitamin A metabolism’ sub-pathway, as well as phosphatidylcholine (p16:0/16:1), 1-linoleoyl glycerol and methyl 4-hydroxybenzoate sulfate. The above mentioned two sub-pathways were enriched for two collections of endophenotypes, cognition and diagnosis, respectively (Supplementary Table S4). Moreover, six metabolites showed associations in males: lysophosphocholine (18:2) (LPC 18:2), sphingomyelin (d38:0), 5-(galactosylhydroxy)-L-Lysine, kynurenate, N-acetylneuraminate and xanthurenate (Supplementary Figure S2-S12).

**Figure 1.**
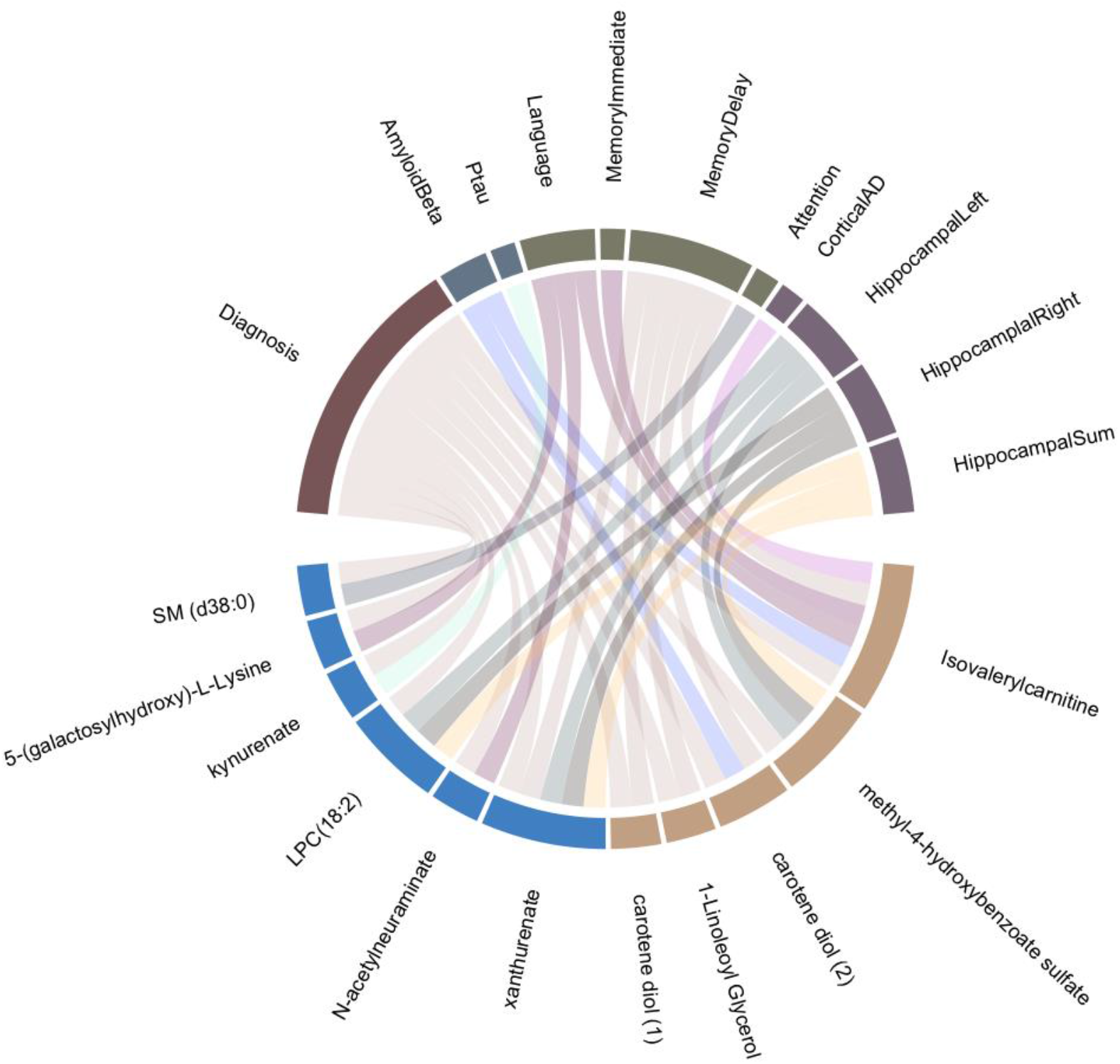
Eleven metabolites showed associations with AD and one other endophenotype. Five plasma metabolites (with brown grid), isovalerylcarnitine, methyl 4-, carotene diol (2), 1-linoleoyl glycerol and carotene diol (1), were significantly associated with diagnosis and the other AD endophenotypes in female group only. Six other metabolites (with blue grid), xanturenate, N-acetylneuraminate, LPC (18:2), kynurenate, 5-(galactosylhydroxy)-L-lysine and SM (d38:0), showed associations with diagnosis and AD variables in male group only.

### 3.3 Interactive effect of *APOE ε4* and sex

After investigating how sex exert metabolite associations with AD endophenotypes, we also performed association analyses for all 540 metabolites with 15 AD endophenotypes from four categories, now stratified by *APOE ε4* status and adjusted for sex (Supplementary methods and results). Supplementary Table S2 summarises the numbers of number of metabolites showed association with each of the AD endophenotypes in the whole cohort and each *APOE ε4* +/- stratum. To investigate potential relationships between sex and *APOE* ε4 status on the metabolomic level, we compared the metabolites identified in either sex group and *APOE* ε4 carrier stratification analyses (Supplementary Table S1 and Table S2). There is no overlap of metabolite that associated with the same AD endophenotype between female/male and *APOE* ε4 + stratum observed at Bonferroni significance thresholds (Supplementary methods and results).

### 3.4 Sex-specific metabolites as diagnostic biomarkers

We further explored sex-specific metabolites, VMA, tryptophan betaine, and kynurenate, as diagnostic biomarkers, that were significant following Bonferroni correction in sex stratified regression analyses. In addition, we tested if these three metabolites were associated to drugs (n = 2, hydroquinone sulfate and salicylate, sample size n = 695), and AD medications (n = 2, acetyl -cholinesterase inhibitors or other AD drugs, sample size = 225). Regression models showed no associations between the three metabolites and drugs at P < 8.17e-04 (data not shown).

In females, a receiver operating characteristic (ROC) curve (bootstrapped) for VMA and tryptophan betaine was built and is presented in Figure 2C (AUC = 0.83 for AD versus controls, SE = 0.029, 95% CI = 0.777 to 0.889). We then examined how well other AD endophenotypes would discriminate AD cases from controls. CSF biomarkers (Aβ, t-tau and p-tau) produced an AUC of 0.89 (SE = 0.025, 95% CI = 0.836 to 0.93), covariates (age at sampling, education years, sampling batch and *APOE ε4* status) produced an AUC of 0.83 (SE = 0.031, 95% CI = 0.763 to 0.886), CSF biomarkers and covariates together produced AUC of 0.92 (SE = 0.019, 95% CI = 0.881 to 0.954). The combination of the two female-specific biomarkers, CSF biomarkers and covariates produced AUC of 0.96 (SE = 0.012, 95% CI = 0.93 to 0.978). Bootstrapping results of standard error and 95% confident interval for AUCs from the ROC curves are listed in Supplementary Table S5.

**Figure 2.**
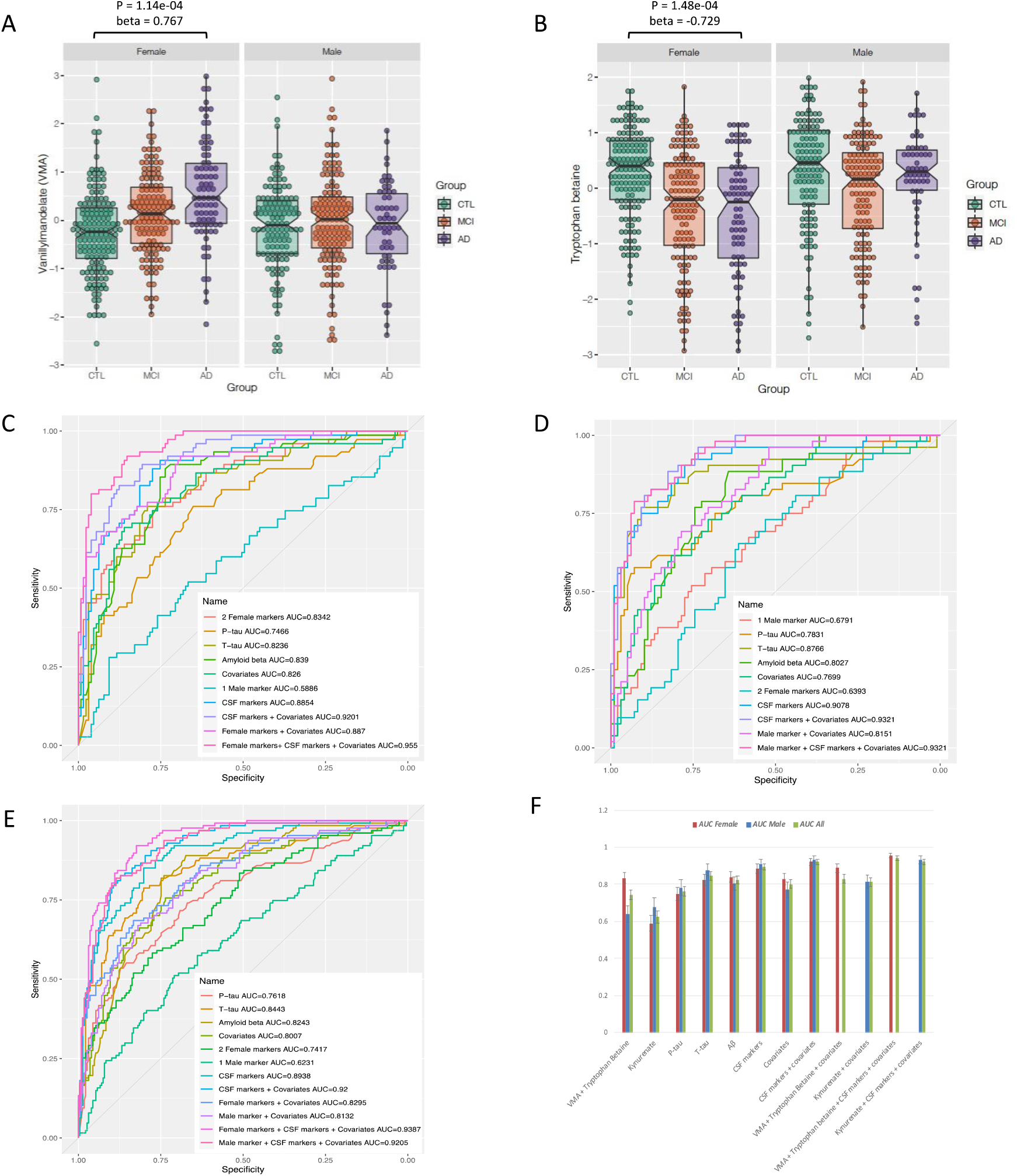
Box plots of two female biomarkers. A) vanillylmandelate and B) tryptophan betaine, the p value (below Bonferroni significance threshold) and the association coefficient (beta) for metabolites in relation to diagnosis (AD/CTL) are shown above the box plots; **ROC graphs, comparing area under curve (AUC) in predictive models using sex-specific biomarkers, CSF biomarkers, covariates, and the combination of them in** C) the female sub-dataset; D) the male sub-dataset; E) the full cohort; **F) Bar plot summarises the AUC of the above mentioned ROC curves together with standard errors after bootstrapping**.

In the male cohort we then built a regression model followed by a ROC curve for kynurenate (Figure 2D). The resulting AUC was 0.68 (SE = 0.046, 95% CI = 0.593 to 0.765). This value was again compared to the CSF biomarkers (AUC = 0.91, SE = 0.026, 95% CI = 0.852 to 0.954), covariates (AUC = 0.77, SE = 0.042, 95% CI = 0.68 to 0.848), both CSF biomarkers & covariates (AUC = 0,932, SE = 0.019, 95% CI = 0.89 to 0.965), and the combination of kynurenate, CSF biomarkers and covariates (AUC = 0.93, SE = 0.02, 95% CI = 0.888 to 0.966). Furthermore, the AUC of CSF biomarkers, covariates, VMA and tryptophan betaine, kynurenate, and combinations of them were explored in the full dataset (males and females combined) (Figure 2E). An increase of AUC from 0.92 to 0.94 was observed when VMA and tryptophan betaine were added to the panel of CSF biomarkers and covariates, while the addition of kynurenate resulted in the same AUC of 0.92 in the full cohort.

Finally, the application of NRI to regression models revealed that, when examining the effect of VMA and tryptophan betaine together in the female sub-dataset, the event NRI, non-event NRI and NRI(p) were 0.04, 0.039, and 0.079 (all above the critical value 0), respectively (Supplementary Figure S13A). However, adding kynurenate to the model in the male sub-dataset yielded nil event NRI, non-event NRI and NRI(p) values (Supplementary Figure S13B). When investigating these sex-specific biomarkers in the full dataset, an NRI(p) of 0.0516 was observed for the two female-specific markers, albeit an NRI(p) of -0.0079 was seen for the male-specific marker (Supplementary Figure S13C & S13D).

## 4 Discussion

In this study, we measured 665 plasma metabolites in 695 participants: 377 females and 318 males. We used a combination of linear and logistic regression analyses as well as pathway enrichment analyses to investigate sex-specific associations between metabolites and AD endophenotypes. For a follow-up investigation, we found that none of the nine metabolites (four primary fatty acid amides, three amino acids, and two lipokines) demonstrating association with AD in our previous study (16) showed interaction with sex in this study which included an additional 102 participants. Overall, we identified ten sub-pathways that showed sex-specific associations in our exploratory pathway analysis (Supplementary methods and results). A number of metabolites demonstrated associations with clinical AD diagnosis and at least one additional AD clinical biomarker. Finally, we report two metabolites, VMA and tryptophan betaine, which had added value to discriminate in females AD cases from controls and demonstrated improved prediction over traditional CSF biomarkers and covariates (AUC of metabolites = 0.834, AUC of CSF markers = 0.884, AUC of CSF markers & covariates = 0.92, total AUC = 0.955).

### 4.1 Metabolites associated with AD and other endophenotypes

When we investigated metabolites associated with diagnosis and at least one other AD endophenotype, five out of 11 metabolites showed significant associations in females, namely isovalerylcarnitine, methyl-4-hydroxybenzoate sulfate, carotene diol (1), carotene diol (2), and 1-lindeoyl glycerol (Figure 1). Among these, isovalerylcarnitine, a short chain acylcarnitine, showed positive associations with cortical thickness in AD regions in females. Moreover, its concentration showed a gradual decrease from cognitively normal group to MCI, and then AD patients. Whilst previous studies have emphasised metabolite links to both sexes, our findings in females are consistent with studies in which sex was not stratified. In these studies, decreased levels of acylcarnitines in schizophrenia and AD were reported (34)(35), although the opposite direction, higher levels of acylcarnitines in subjects with AD have also been observed (36). Carotene diols are unoxygenated carotenoids involved in vitamin A metabolism and we observed decreased levels in AD patients, while other studies have also shown that beta-carotene might protect against dementia (37).

Six metabolites associated with AD and at least one other AD endophenotype in males, namely xanthurenate, N-acetylneuraminate, LPC (18:2), kynurenate, 5-(galactosylhydroxy)-L-lysine, and SM(d38:0). While our study focuses on novel metabolites with association to male sex, links to cognition and AD have been previously reported. Xanthurenate and kynurenate are formed in the kynurenine pathway by tryptophan degradation. They may play a physiological role in attentional and cognitive processes, as well as act as a potential trait marker for schizophrenia (38). The importance of lipid dynamics has been studied as an area of investigation in AD (39). In this regard, we observed significantly increased levels of LPC (18:2) – a glycerophospholipid - in males, although decreased plasma LPC (18:2) levels have been reported in a non-targeted AD study (40).

### 4.2 Sex-specific biomarkers

In this study, we found three metabolites, VMA, tryptophan betaine and kynurenine in the tyrosine and tryptophan pathways, to be associated with AD in two sex strata. Kaddurah-Daouk *et al*. found that the level of VMA was elevated in AD versus controls in CSF (41), in accordance with our findings. VMA is an end product of catecholamine metabolism in the tyrosine-dopamine pathway (Supplementary Figure S14A) metabolized by catechol-O-methytransferase (COMT) and monoamine oxidase (MAO) (41). Therefore, the observed increase in positive association with VMA could be the result of the upregulation COMT and/or MAO in female AD patients. Indeed, oestrogen turnover via COMT has been implicated in AD pathogenesis in ApoE-dependent manner (42). In addition, the activation of MAO-B, the alternative pathway to VMA metabolism, has been demonstrated in brains with AD (43).

Tryptophan betaine was the second female-specific AD marker in this study. An important finding of this study is that metabolites that can act via the gut-brain axis showed sex-specific associations. Tryptophan betaine is found in high fibre plants, and it is also linked to microbiota in fibre-enriched diets (44). Koistinen *et al*. found that tryptophan betaine correlated negatively with several bacterial taxa which have earlier been associated with adverse effects in the gut and overall health in an in vitro model of the human gastrointestinal system (44). It is an indole alkaloid composed of tryptophan and three methyls (Supplementary Figure S14B) and has been found to have neurological and glucose-lowering effects in rodents (45). Interestingly, it has been also been reported as sleep-inducing and anti-inflammatory compound when tested in animal models (45,46). Furthermore, in females, our pathway analyses revealed enrichment in the tryptophan pathway for AD diagnosis (Supplementary methods and results).

Another metabolite in the same pathway and that was decreased in male AD patients specifically was kynurenate. It is produced via a secondary branch of the kynurenine pathway through tryptophan degradation (Supplementary Figure S14B). This is in agreement with studies with no sex-specific analyses, where kynurenine concentrations were decreased in AD plasma/serum (36)(47). In the past it has been reported as an endogenous excitatory amino acid receptor antagonist, as well as a neuroprotective compound (48). Tryptophan is the parent metabolite in both the serotonin and kynurenine pathways. Kynurenate can lead to attenuated glutamate neurotransmission by antagonism of glutamate receptors and release, which consequently decreases glutamate-evoked dopamine release in the striatum (49).

Both pathways, tyrosine-dopamine and tryptophan, have been reported to be linked to inflammation in psychiatric disease (50). The predictive abilities of the two female- and one male-specific biomarkers were initially evaluated by calculating the AUC, and further assessed by NRI when compared to existing baseline models. NRI(p) of 0.079 and 0.052 were observed for the two female-specific markers in the female dataset and the full dataset (Supplementary Figure S13A & S13C), respectively. With the baseline AUC over 0.9, it has been reported that the effect size (Cohen’s D) of adding the female-specific markers could reach 0.8 (33). The increased AUC values, together with positive NRI, support the hypothesis that female-specific AD markers add value to precision medicine (31).

### 4.3 Limitations and conclusions

A limitation of this study is that, although it is relatively large, the number of participants is still small particularly after sex-stratification. Also, results of this study should be validated in an independent cohort. However, to avoid potential overfitting all AUCs from ROC curves were bootstrapped 1000 times with 95% CIs and standard errors presented. In addition, an important finding of this study is the gut-brain axis link shown by the tryptophan betaine and VMA pathways in females, however no causal implications can be drawn from our study. Additional limitations include survival and selection bias inherent to observational studies. In summary, our study reports sex-specific metabolic differences in AD and highlights two metabolites, both involved in neurotransmission pathways as potential female-specific biomarkers in AD. This is the first report that highlights female-specific and modifiable metabolites associated with AD, paving the way to personalised treatment.

## Data Availability

Data used in preparation of this article were obtained from the European Medical Information Framework for Alzheimer's Disease Multimodal Biomarker Discovery (EMIF-AD MBD) cohort.

## Acknowledgements

The authors thank the participants and families who took part in this research. The authors would also like to thank all people involved in data and sample collection and/or logistics across the different centres.

## Appendix

List of collaborators in European Medical Information Framework (EMIF) consortium.

| Organisations | Partner type | Country |
| --- | --- | --- |
| Aarhus Universitetshospital | Academia | Denmark |
| Agenzia Regionale di Sanità | Academia | Italy |
| Alzheimer Europe | Patient Organisation | Luxembourg |
| Amgen | EFPIA | UK |
| Boehringer Ingelheim International GmbH | EFPIA | Germany |
| Cambridge Cognition Ltd | SME | UK |
| Concentris Research Management GmbH | SME | Germany |
| Custodix NV | SME | Belgium |
| Electrophoretics Ltd | SME | UK |
| Erasmus Universitair Medisch Centrum Rotterdam | Academia | The Netherlands |
| European Institute for Health Records | Academia | Belgium |
| European Molecular Biology Laboratory | Academia | Germany |
| F. Hoffmann-La Roche AG | EFPIA | Switzerland |
| Fondazione PENTA - for the Treatment and Care of Children with HIV-ONLUS | Academia | Italy |
| Genomedics S.R.L. | Academia | Italy |
| GlaxoSmithKline Research and Development Ltd | EFPIA | UK |
| Göteborgs Universitet | Academia | Sweden |
| Helsingin Yliopisto | Academia | Finland |
| IDIAP | Academia | Spain |
| Institut National de la Santé et de la Recherche Médicale | Academia | France |
| Itä-Suomen Yliopisto | Academia | Finland |
| Janssen Pharmaceutica NV | EFPIA | Belgium |
| Karolinska Institutet | Academia | Sweden |
| King's College London | Academia | UK |
| Leibniz Institute for Prevention, Research and Epidemiology - BIPS GmbH | Academia | Germany |
| Maastricht University | Academia | The Netherlands |
| MAAT France | SME | France |
| Merck | EFPIA | Germany |
| Novo Nordisk A/S | EFPIA | Denmark |
| Pedianet | SME | Italy |
| Pfizer | EFPIA | UK |
| Pharmo Cooperatie UA | SME | The Netherlands |
| Provincia Lombardo-Veneta -<br>Ordine Ospedaliero di San<br>Giovanni di Dio—<br>Fatebenefratelli | Academia | Italy |
| Servier | EFPIA | France |
| Synapse Research Management<br>Partners S.L. | SME | Spain |
| Tartu Ulikool | Academia | Estonia |
| Teknologian tutkimuskeskus<br>VTT | Academia | Finland |
| UCB Pharma SA | EFPIA | Belgium |
| Universidade de Aveiro | Academia | Portugal |
| Universitat Pompeu Fabra | Academia | Spain |
| Universiteit Antwerpen | Academia | Belgium |
| University College London | Academia | UK |
| University of Copenhagen | Academia | Denmark |
| University of Exeter | Academia | UK |
| University of Glasgow | Academia | UK |
| University of Leicester | Academia | UK |
| University of Manchester | Academia | UK |
| University of Oxford | Academia | UK |
| University Pierre et Marie<br>Curie | Academia | France |
| Università degli Studi di Pisa | Academia | Italy |
| Universität Leipzig | Academia | Germany |
| Universität zu Lübeck<br>(University of Lübeck) | Academia | Germany |
| Universitätsklinikum Erlangen,<br>Anstalt des Öffentlichen<br>Rechts | Academia | Germany |
| Vestische Kinder - und<br>Jugendklinik Datteln | Academia | Germany |
| VIB | Academia | Belgium |
| VU University Medical Center | Academia | The Netherlands |

