## Supplementary document for "Sex-specific metabolic pathways associate with Alzheimer’s Disease (AD) endophenotypes in the European Medical Information Framework for AD Multimodal Biomarker Discovery cohort"

### Supplementary methods and results

#### Pathway enrichment analysis

The hypergeometric test was performed to identify pathways that are enriched in females and males in collections of AD endophenotypes (diagnosis, CSF markers, cognition, and brain MRI) for metabolites showing associations at the nominal significance level in sex stratified analyses (1)(2). P-values were adjusted for false discovery rate (FDR) at a corrected P<0.05.

Each known metabolite was classified into one of the following eight “super-pathways”: ‘Amino acid’, ‘Carbohydrate’, ‘Cofactors and vitamins’ ‘Energy’, ‘Lipid’, ‘Nucleotide’, ‘Peptide’, and ‘Xenobiotics’ by *Metabolon* based on the Kyoto Encyclopaedia of Genes and Genomes (KEGG) database. Each super-pathway was further subdivided into two or more “sub-pathways”. The original 60 sub-pathways pre-defined by *Metabolon* for all 665 metabolites were employed as reference. Over-representation analyses using the hypergeometric test were performed for the metabolites associated in the sex-stratified analyses with the collections of endophenotypes in females and males respectively. Those containing less than 5 metabolites were excluded, resulting in 34 sub-pathways (10 sub-pathways for diagnosis, 10 for CSF biomarkers, 27 for cognition outcomes and 16 for brain MRI measurements) in the female dataset and 39 sub-pathways (10 sub-pathways for diagnosis, 10 for CSF biomarkers, 26 for cognition outcomes and 18 for brain MRI measurements) in the male dataset. It was revealed that ‘vitamin A metabolism’ (q = 5.7e-04) and ‘tryptophan metabolism’ sub-pathways (q = 2.4e-02) were enriched for diagnosis in the female group. Three additional sub-pathways were associated with cognition outcomes and four sub-pathways with brain MRI measurements in females (Supplementary Table S2), including ‘acyl carnitine fatty acid metabolism’ (q = 8.4e-04), ‘monohydroxy fatty acid’ (q = 1.7e-02), ‘leucine, isoleucine and valine metabolism’ (q = 4.1e-02), ‘sphingolipid metabolism’ (q = 3.4e-

02), ‘phosphatidylinositol (PI)’ (q = 1.9e-02), ‘primary bile acid metabolism’ (q = 3.4e-02), and ‘plasmalogen’ (q = 3.4e-04). Ceramides were enriched for both cognition (q = 6.6e-04) and brain MRI measurements (q = 1.7e-04) in males.

The hypergeometric analysis highlighted nine sub-pathways in females and one in males. In females, both vitamin A metabolism (3) and tryptophan metabolism pathways (4) were correlated with diagnosis. The other important sub-pathways in females included acyl carnitine metabolism, phosphatidylinositol, as well as leucine, isoleucine and valine (5) which have been reported as combating oxidative stress, AD pathology or cognition. It has been demonstrated in other studies that the metabolism of some acyl-carnitines is finely connected among different diagnostic groups, as well as with age and sex (6). In males, the ceramides sub-pathway, which has been widely reported to play a role in AD and other neurodegenerative disorders was enriched (7).

**Association analyses including *APOE ε4* as stratification variable**

Studies have reported that *APOE ε4* genotype may exert AD risk in a sex-dependent manner, which has a greater influence on female *APOE ε4* carrier (8–11).To investigate potential influence of *APOE ε4* status on the metabolomic level, we also performed association analyses with all 15 AD endophenotypes from four categories, now stratified by *APOE ε4* status and adjusted for sex. We found that 118 metabolites showed asscociation with AD endophenotypes at nominal significance level in the *APOE ε4* carrier stratum, while 116 metabolites were predominant in the non-carrier stratum (supplementary table S2). Similarly, there were 22 and 18 metabolites associated with AD endophenotypes in *APOE ε4* + and *APOE ε4* – stratum at Bonferroni significance threshold ($P < 8.17e-04$), respectively.

After comparing the metabolites identified in either sex group and *APOE* ε4 carrier stratification analyses (Supplementary Table S1 and Table S2). It was noticed that one metabolite, 2-Hydroxyisocaproic acid, showed association with immediate memory at nominal level in both female and *APOE* ε4 carrier strata (Supplementary Table S3 & Figure S15F). Additionally, the overlaps of metabolites between male statum and *APOE* ε4 carrier stratum at nominal level include LPC (18:2) and methyl glucopyranoside (alpha + beta) (associated with AD), cystathionine (associated with attention), Cer(d18:1/16:0) and GlcCer(d18:1/16:0) (associated with visuoconstruction) (Supplementary Table S3 & Figure S15A-E). Finally, when we investigated further for the cross-over overlapping metabolites that showed association with the the same endophenotype from sex strata at Bonferroni threshold and *APOE* ε4 strata at nominal level, as well as those from *APOE* ε4 starta at Bonferroni threshold and sex starta at nominal level, LPC (18:2) was found to be the only metabolite that associated with AD in males (P = 1.11e-03) and *APOE* ε4 carriers (P = 3.92e-04).

### Supplementary Tables

**Table S1. Number of metabolites showed association with each AD endophenotypes in the whole cohort and each sex stratum.**

| *Category* | *Outcomes* | *All(*sex) p<0.05* | | *Female p<0.05* | | *Male p<0.05* | | *Female p<8.17e-04* | *Male p<8.17e-04* |
|---|---|---|---|---|---|---|---|---|---|
| Diagnosis | AD/CTL | 30 | | 15 | | 13 | | 2 | 1 |
| CSF markers | P-tau | 11 | (38) | 5 | (15) | 5 | (13) | | |
| | T-tau | 2 | | 6 | | 1 | | | |
| | Amyloid-beta | 26 | | 6 | | 7 | | | |
| Cognition | Attention | 13 | (152) | 7 | (66) | 7 | (60) | | |
| | Executive function | 48 | | 16 | | 7 | | | |
| | Language | 27 | | 6 | | 19 | | | |
| | Memory Delay | 27 | | 18 | | 8 | | | |
| | Memory Immediate | 26 | | 19 | | 6 | | | |
| | Visuoconstruction | 34 | | 10 | | 17 | | | |
| Brain MRI | Hippocampal Left | 23 | (79) | 8 | (35) | 10 | (33) | | |
| | Hippocampal Right | 26 | | 6 | | 12 | | | |
| | Hippocampal Sum | 22 | | 6 | | 10 | | | |
| | Cortical thickness WB | 28 | | 9 | | 8 | | | |
| | Cortical thickness AD | 43 | | 21 | | 14 | | | |
| Total | | **230** | | **116** | | **100** | | | |

WB, whole brain; AD, Alzheimer's disease; CTL, control.

**Table S2. Number of metabolites showed association with AD endophenotypes in the whole cohort and each *APOE* ε4 +/- stratum.**

| *Category* | *Outcomes* | *All(*APOE ε4 +/-) p<0.05* | *APOE ε4 + p<0.05* | | *APOE ε4 - p<0.05* | | *APOE ε4 + p<8.17e-04* | | *APOE ε4 - p<8.17e-04* | |
|---|---|---|---|---|---|---|---|---|---|---|
| Diagnosis | AD/CTL | 42 | 23 | | 13 | | 7 | | 1 | |
| CSF markers | P-tau | 65 | 25 | | 34 | | 1 | | 8 | |
| | T-tau | 66 | 24 | (40) | 37 | (60) | - | (2) | 10 | (16) |
| | Amyloid-beta | 31 | 6 | | 12 | | 1 | | 3 | |
| Cognition | Attention | 35 | 16 | | 12 | | 1 | | 2 | |
| | Executive function | 21 | 8 | | 8 | | - | | 1 | |
| | Language | 9 | 3 | | 3 | | 2 | | 1 | |
| | Memory Delay | 44 | 16 | (62) | 8 | (41) | 3 | (8) | - | (5) |
| | Memory Immediate | 32 | 13 | | 10 | | 2 | | 2 | |
| | Visuoconstruction | 43 | 11 | | 4 | | - | | - | |
| Brain MRI | Hippocampal Left | 18 | 8 | | 4 | | 2 | | - | |
| | Hippocampal Right | 12 | 3 | | 5 | | 1 | | - | |
| | Hippocampal Sum | 15 | 7 | (15) | 5 | (14) | 2 | (2) | 1 | |
| | Cortical thickness WB | 14 | 5 | | 3 | | - | | - | |
| | Cortical thickness AD | 17 | 4 | | 5 | | - | | - | |
| Total | | | **118** | | **116** | | **18** | | **22** | |

WB, whole brain; AD, Alzheimer's disease; CTL, control.

**Table S3**. **Overlap of of metabolites showed association with AD endophenotypes in the stratification analyses between female/male group and *APOE* ε4 + group at nominal significance levels ($p < 0.05$).**

| Sex | AD endophenotypes | *APOE ε*4 + group |
|---|---|---|
| Male | Diagnosis | LPC (18:2) |
| | | methyl glucopyranoside (alpha + beta) |
| | Attention | Cystathionine |
| | Visuoconstruction | Cer(d18:1/16:0) |
| | | GlcCer(d18:1/16:0) |
| Female | Memory Immediate | 2-Hydroxyisocaproic acid |

**Table S4**. **The associations of the ten sub-pathways with four collections of AD endophenotypes in both sex strata.**

| | | Pathways | P-value | q-value |
|---|---|---|---|---|
| Female | Diagnosis | Vitamin A metabolism | 5.73e-05 | 5.7e-04 |
| | | Tryptophan metabolism | 4.71e-03 | 2.4e-02 |
| | Cognition | Acyl Carnitine metabolism | 3.10e-05 | 8.4e-04 |
| | | Monohydroxy fatty acid | 1.26e-03 | 1.7e-02 |
| | | Leucine, isoleucine and valine metabolism | 4.58e-03 | 4.1e-02 |
| | MRI | Phosphatidylinositol metabolism | 1.17e-03 | 1.9e-02 |
| | | Sphingolipid metabolism | 4.28e-03 | 3.4e-02 |
| | | Primary bile acid metabolism | 6.29e-03 | 3.4e-02 |
| | | Plasmalogen | 8.40e-03 | 3.4e-02 |
| Male | Cognition | Ceramides | 2.54e-05 | 6.6e-04 |
| | MRI | Ceramides | 9.60e-06 | 1.7e-04 |

**Table S5. Bootstrapping results of standard error (SE) and 95% confident interval for AUCs from ROC curves.**

| | AUC | | | SE | | | 95% confidence interval | | |
|---|---|---|---|---|---|---|---|---|---|
| | Female | Male | All | Female | Male | All | Female | Male | All |
| VMA + Tryptophan Betaine | 0.834 | 0.639 | 0.742 | 0.029 | 0.046 | 0.027 | 0.777 – 0.889 | 0.546 – 0.733 | 0.687 – 0.795 |
| Kynurenate | 0.589 | 0.679 | 0.623 | 0.043 | 0.046 | 0.031 | 0.505 – 0.672 | 0.593 – 0.765 | 0.562 – 0.682 |
| P-tau | 0.748 | 0.783 | 0.762 | 0.037 | 0.042 | 0.027 | 0.672 – 0.816 | 0.697 – 0.862 | 0.707 – 0.813 |
| T-tau | 0.824 | 0.877 | 0.845 | 0.031 | 0.035 | 0.023 | 0.756 – 0.881 | 0.805 – 0.942 | 0.798 – 0.887 |
| Aβ | 0.839 | 0.803 | 0.824 | 0.029 | 0.035 | 0.022 | 0.775 – 0.892 | 0.734 – 0.865 | 0.781 – 0.866 |
| CSF markers | 0.885 | 0.908 | 0.894 | 0.025 | 0.026 | 0.018 | 0.836 – 0.93 | 0.852 – 0.954 | 0.858 – 0.926 |
| Covariates | 0.826 | 0.77 | 0.801 | 0.031 | 0.042 | 0.024 | 0.763 – 0.886 | 0.68 – 0.848 | 0.752 – 0.846 |
| CSF markers + covariates | 0.92 | 0.932 | 0.921 | 0.019 | 0.019 | 0.014 | 0.881 – 0.954 | 0.89 – 0.965 | 0.893 – 0.948 |
| VMA + Tryptophan Betaine + covariates | 0.887 | - | 0.83 | 0.024 | - | 0.023 | 0.836 – 0.931 | - | 0.784 – 0.874 |
| Kynurenate + covariates | - | 0.815 | 0.813 | - | 0.035 | 0.023 | - | 0.742 – 0.878 | 0.766 – 0.854 |
| VMA + Tryptophan betaine + CSF markers + covariates | 0.955 | - | 0.939 | 0.012 | - | 0.012 | 0.93 – 0.978 | - | 0.914 – 0.962 |
| Kynurenate + CSF markers + covariates | - | 0.932 | 0.921 | - | 0.02 | 0.014 | - | 0.888 – 0.966 | 0.893 – 0.948 |

Covariates include age at sampling, education years, sampling batch and *APOE ε4* status; CSF makers include p-tau, t-tau and Aβ; AUC, area under curve; ROC, receiver operating characteristic; VMA, vanillymadelate, Aβ, Amyloid beta.

### Supplementary Figures

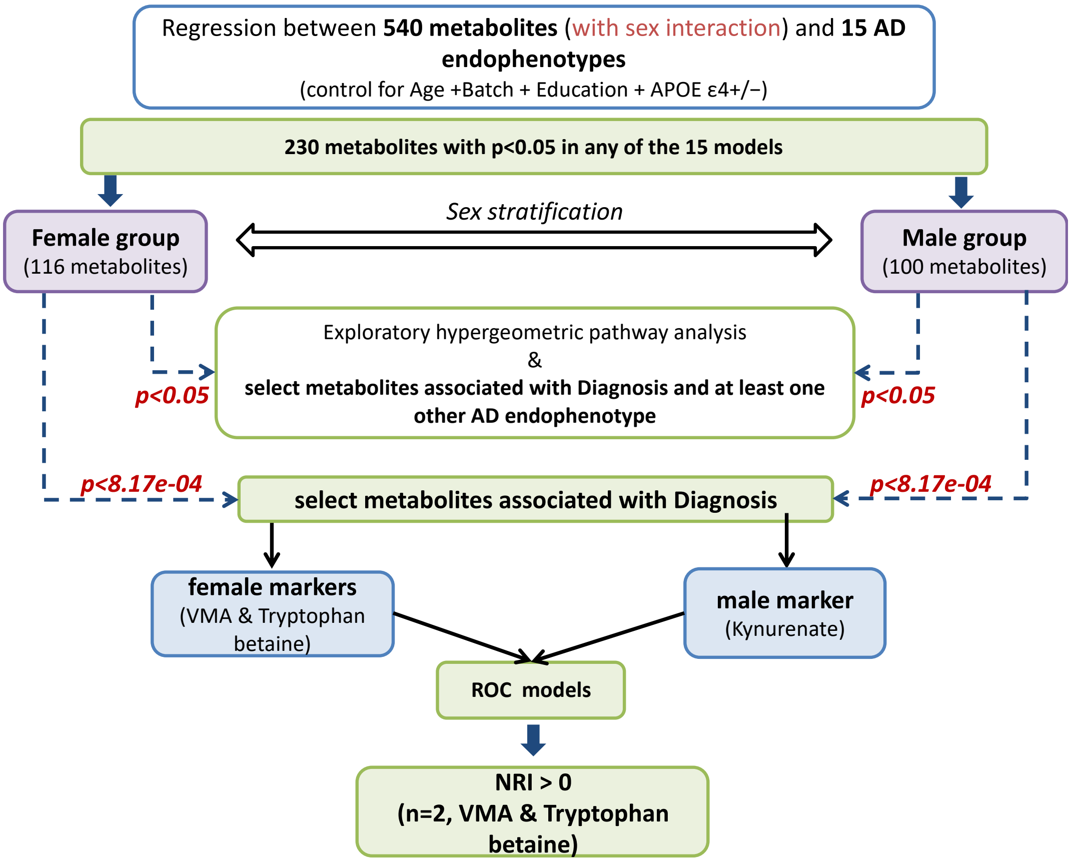


**Figure S1. Study workflow.** The associations between 540 plasma metabolites with a sex interaction term and 15 AD endophenotypes were investigated using linear/logistic regression. These models were repeated for 230 significant metabolites that showed interactions with sex. Metabolites that i) passed the P<0.05 threshold and correlated with any of 15 AD endophenotypes were further employed for exploratory hypergeometric pathway analysis in four categories of AD endophenotypes; ii) passed the P<0.05 threshold and correlated with both diagnosis as well as any other endophenotype were investigated; iii) passed the p<8.17e-04 threshold in each sex stratum and showed association with diagnosis (AD/CTL) were selected for regession models followed by receiver operating characteristic (ROC) curves to test their prediction abilities in AD, which was further validated using net reclassification index (NRI).

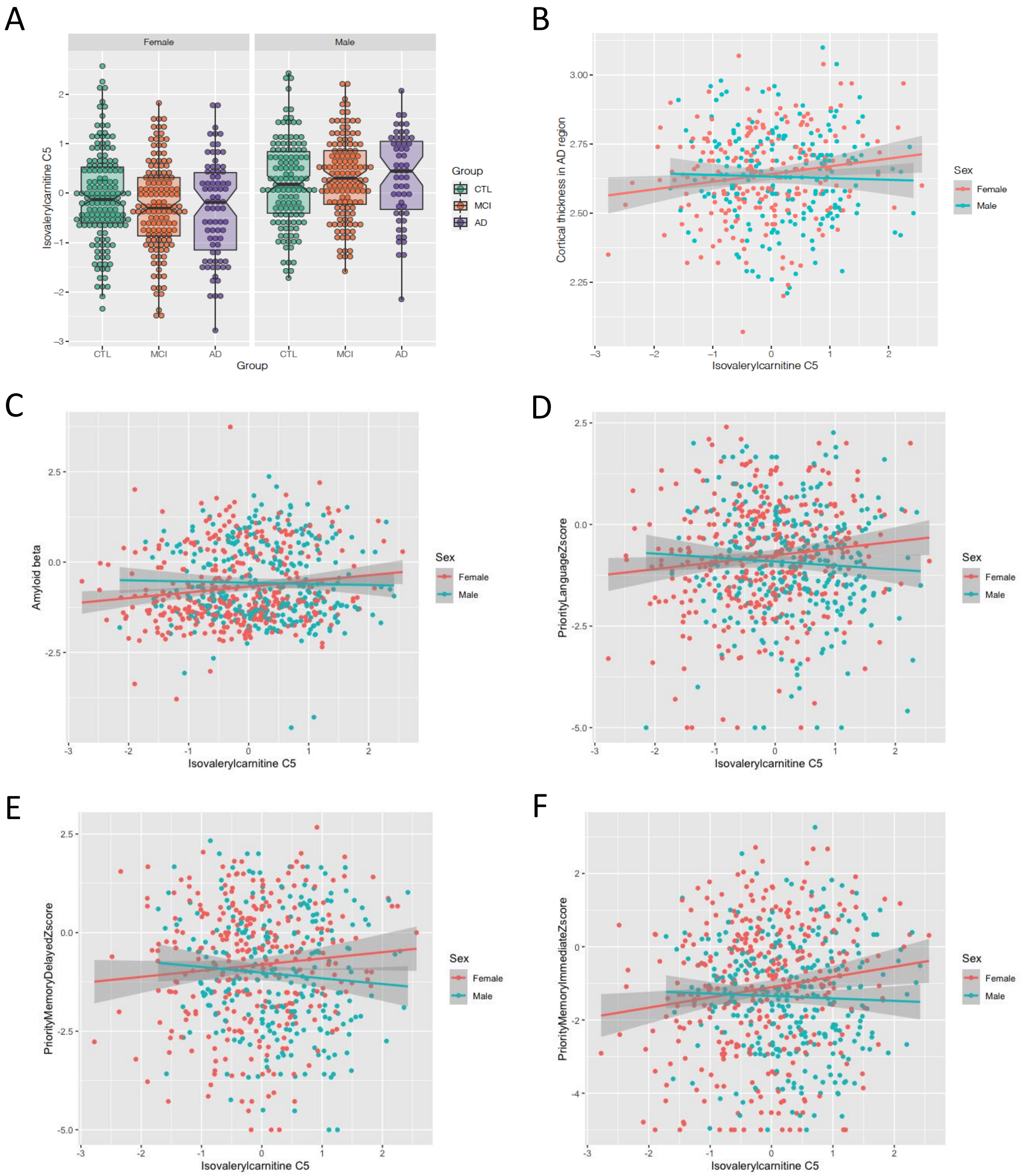


**Figure S2**. Isovalerylcarnitine C5 showed significant association in the female group with A) diagnosis (AD/CTL) ($\beta = -0.46$, 95% CI = -0.82 to -0.11, P = 1.1e-02); B) cortical thickness in AD region ($\beta = 0.031$, 95% CI = 0.0085 to 0.053, P = 7.3e-03); C) amyloid-beta ($\beta = 0.17$, 95% CI = 0.065 to 0.27, P = 1.3e-03); D) language z-score ($\beta = 0.18$, 95% CI = 0.043 to 0.33, P = 1.1e-02); E) memory delay z-score ($\beta = 0.19$, 95% CI = 0.0052 to 0.38, P = 4.5e-02)and F) memory immediate z-score ($\beta = 0.29$, 95% CI = 0.078 to 0.50, P = 7.5e-03).

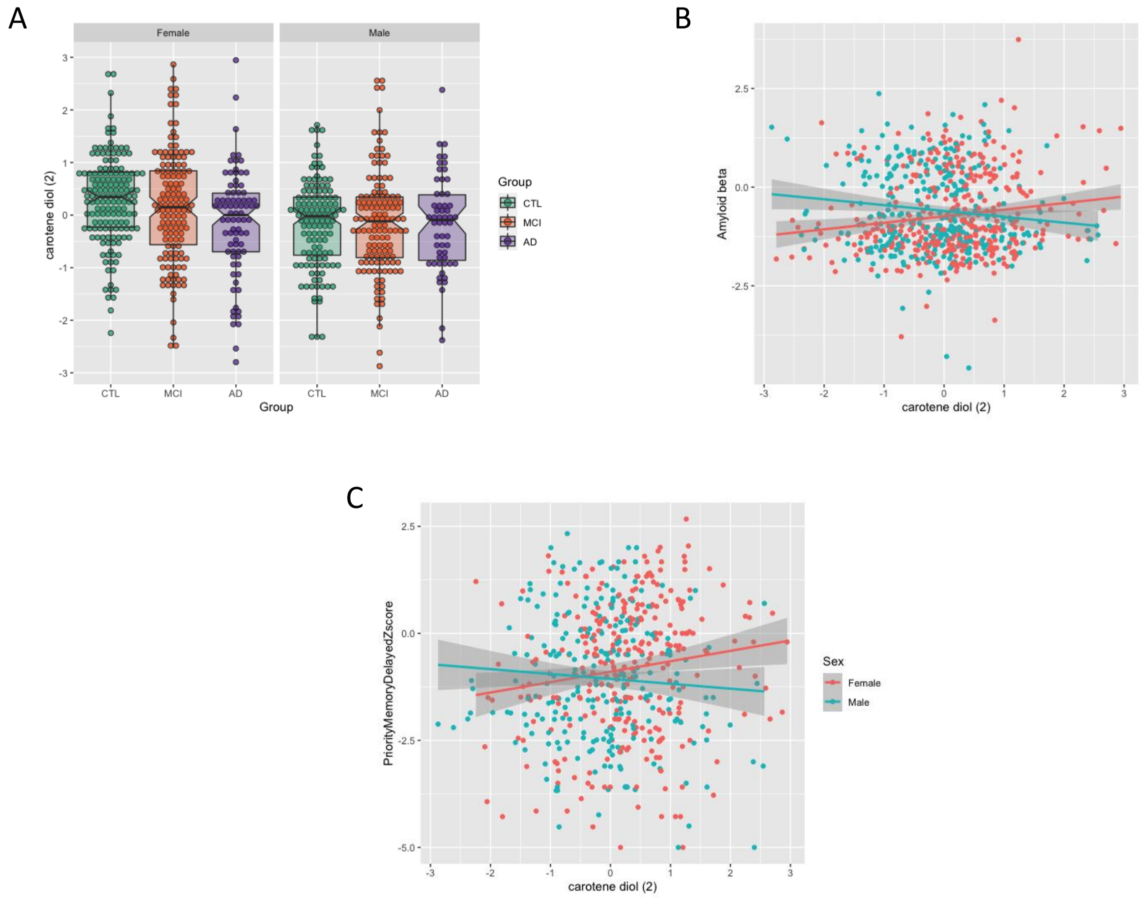


**Figure S3**. Carotene diol (2) showed significant association in the female group with A) diagnosis (AD/CTL) (β = -0.56, 95% CI = -0.93 to -0.22, P = 1.94e-03); B) amyloid-beta (β = 0.13, 95% CI = 0.033 to 0.23, P = 9.1e-03) and C) memory delay z-score (β = 0.022, 95% CI = 0.036 to 0.40, P = 1.9e-02).

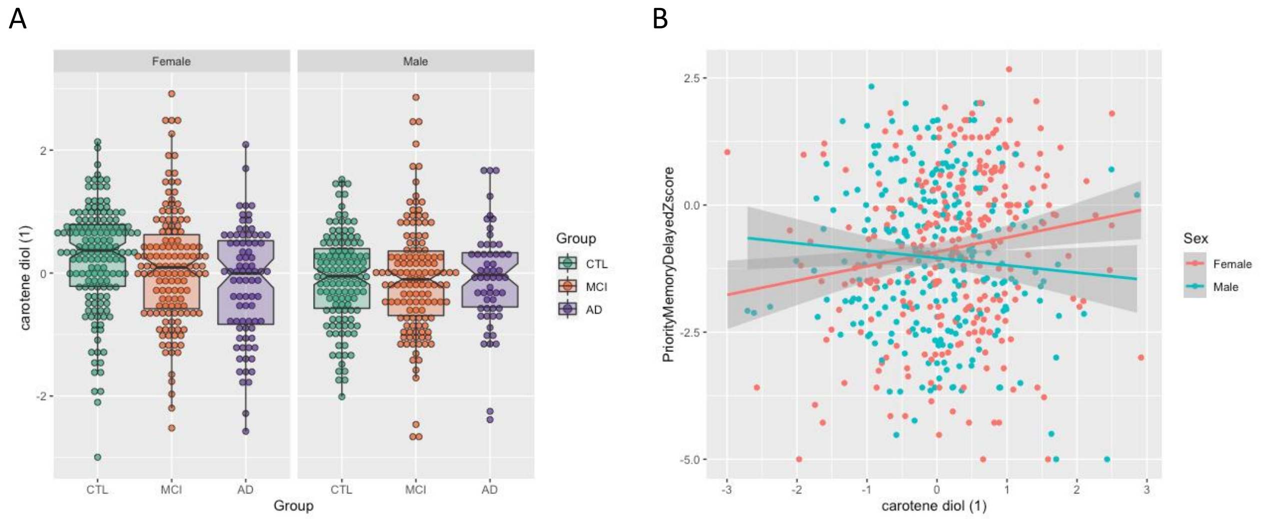


**Figure S4**. Carotene diol (1) showed significant association in the female group with A) diagnosis (AD/CTL) (β = -0.58, 95% CI = -0.96 to -0.22, P = 1.95e-03) and B) memory delay z-score (β = 0.21, 95% CI = 0.021 to 0.40, P = 3.0e-02).

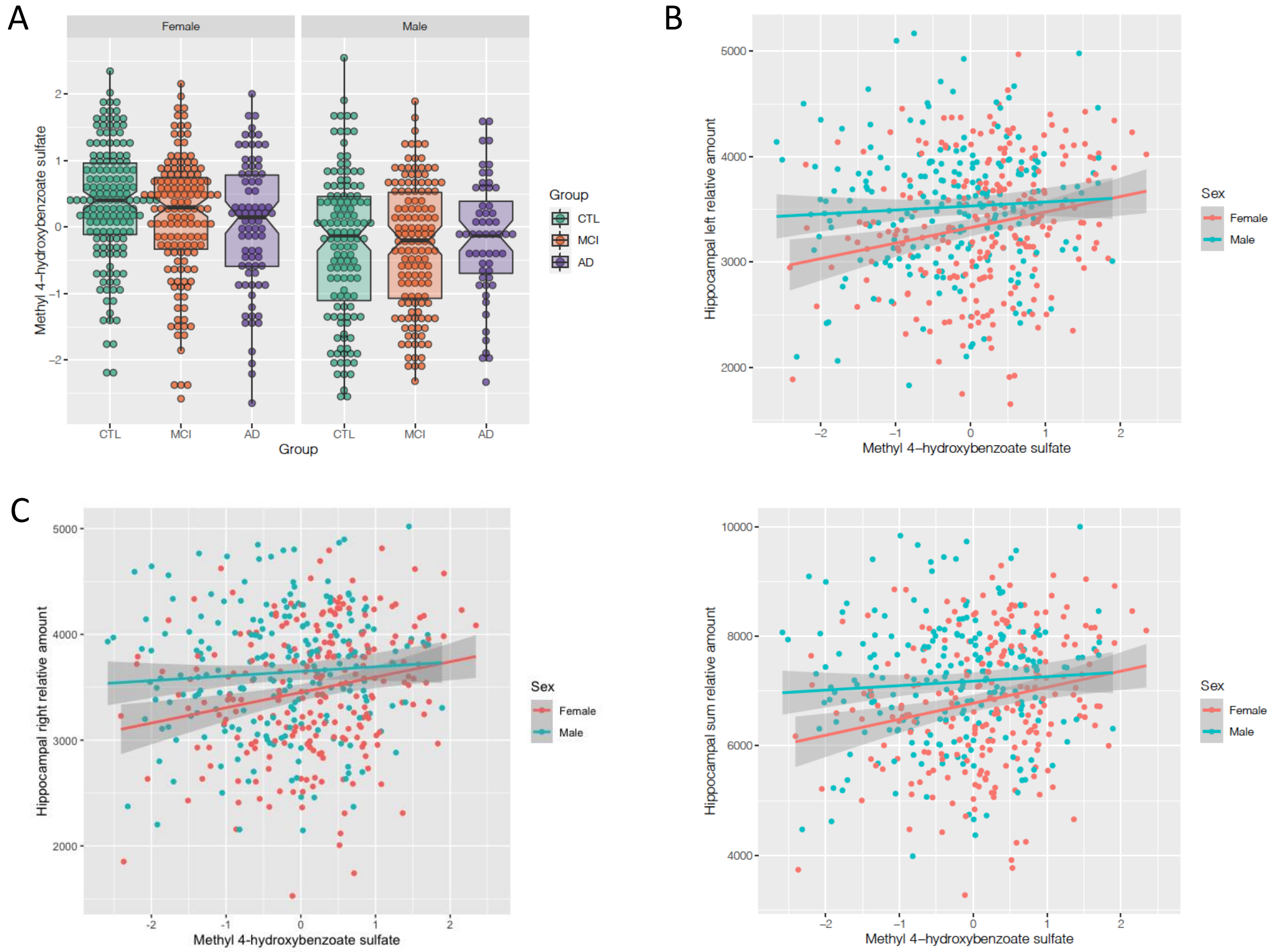


**Figure S5**. Methyl 4-hydroxybenzoate sulfate showed significant association in the female group with A) diagnosis (AD/CTL) (β = -0.41, 95% CI = -0.76 to -0.075, P = 1.8e-02); B) hippocampal left relative amount (β = 131.99, 95% CI = 51.86 to 212.13, P = 1.4e-03); C) hippocampal right relative amount (β = 123.74, 95% CI = 45.80 to 201.69, P = 2.1e-03) and D) hippocampal total relative amount (β = 255.73, 95% CI = 104.46 to 407.00, P = 1.1e-03).

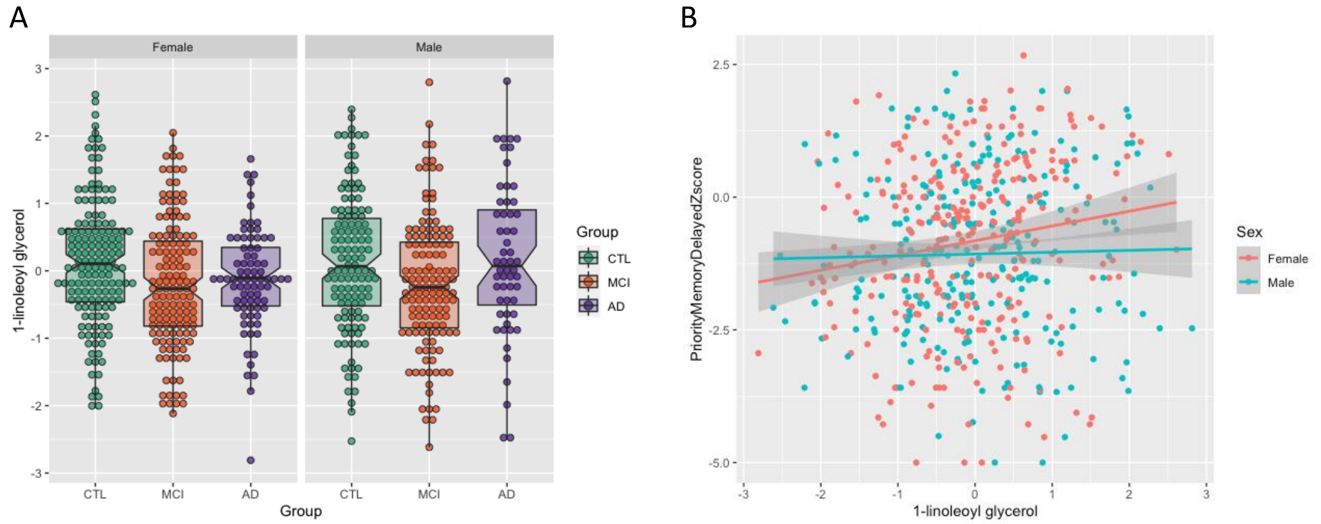


**Figure S6**. 1-linoleoyl glycerol showed significant association in the female group with A) diagnosis (AD/CTL) (β = -0.57, 95% CI = -0.98 to -0.18, P = 4.7e-03) and B) memory delay z-score (β = 0.27, 95% CI = 0.09 to 0.46, P = 3.7e-03).

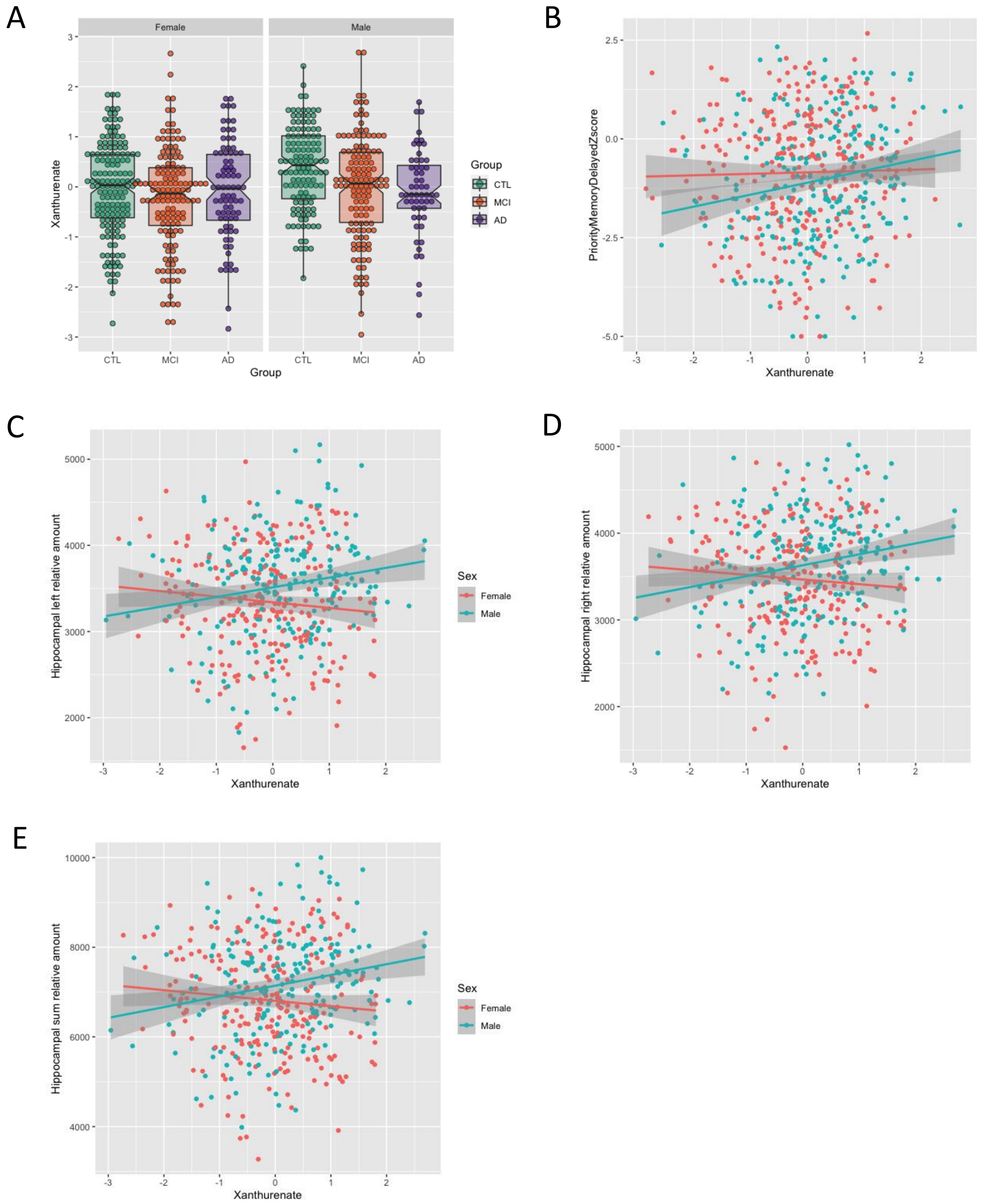


**Figure S7**. Xanthurenate showed significant association in the male group with A) diagnosis (AD/CTL) (β = -0.73, 95% CI = -1.18 to -0.31, P = 1.02e-03); B) amyloid-beta (β = 0.14, 95% CI = 0.026 to 0.25, P = 1.62e-02); C) hippocampal left relative amount (β = 83.28, 95% CI = 8.83 to 157.72, P = 2.9e-02); D) hippocampal right relative amount (β = 98.28, 95% CI = 25.45 to 171.11, P = 8.8e-03) and E) hippocampal total relative amount (β = 181.57, 95% CI = 41.18 to 321.95, P = 1.2e-02).

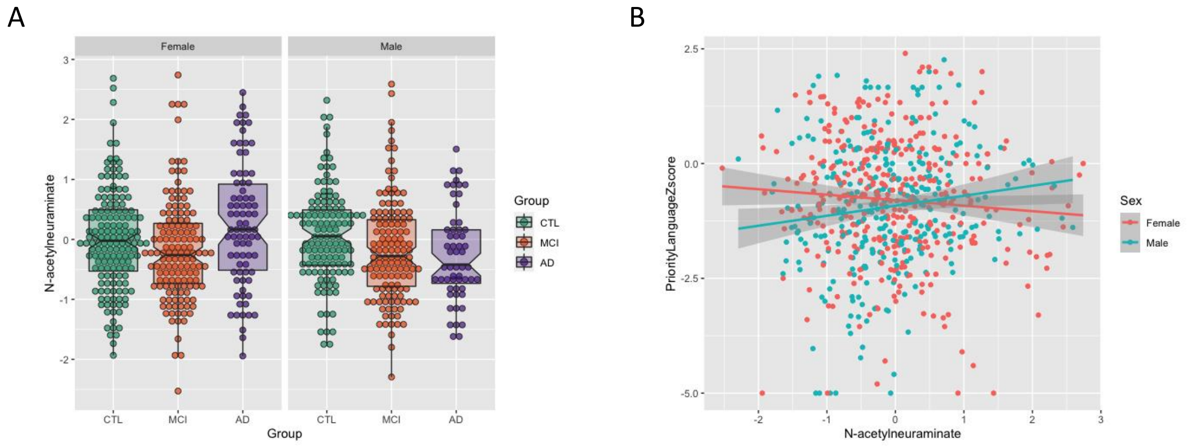


**Figure S8.** N-acetylneuraminate showed significant association in the male group with A) diagnosis (AD/CTL) (β = -0.64, 95% CI = -1.13 to -0.18, P = 8.03e-03) and B) language z-score (β = 0.23, 95% CI = 0.046 to 0.42, P = 1.5e-02).

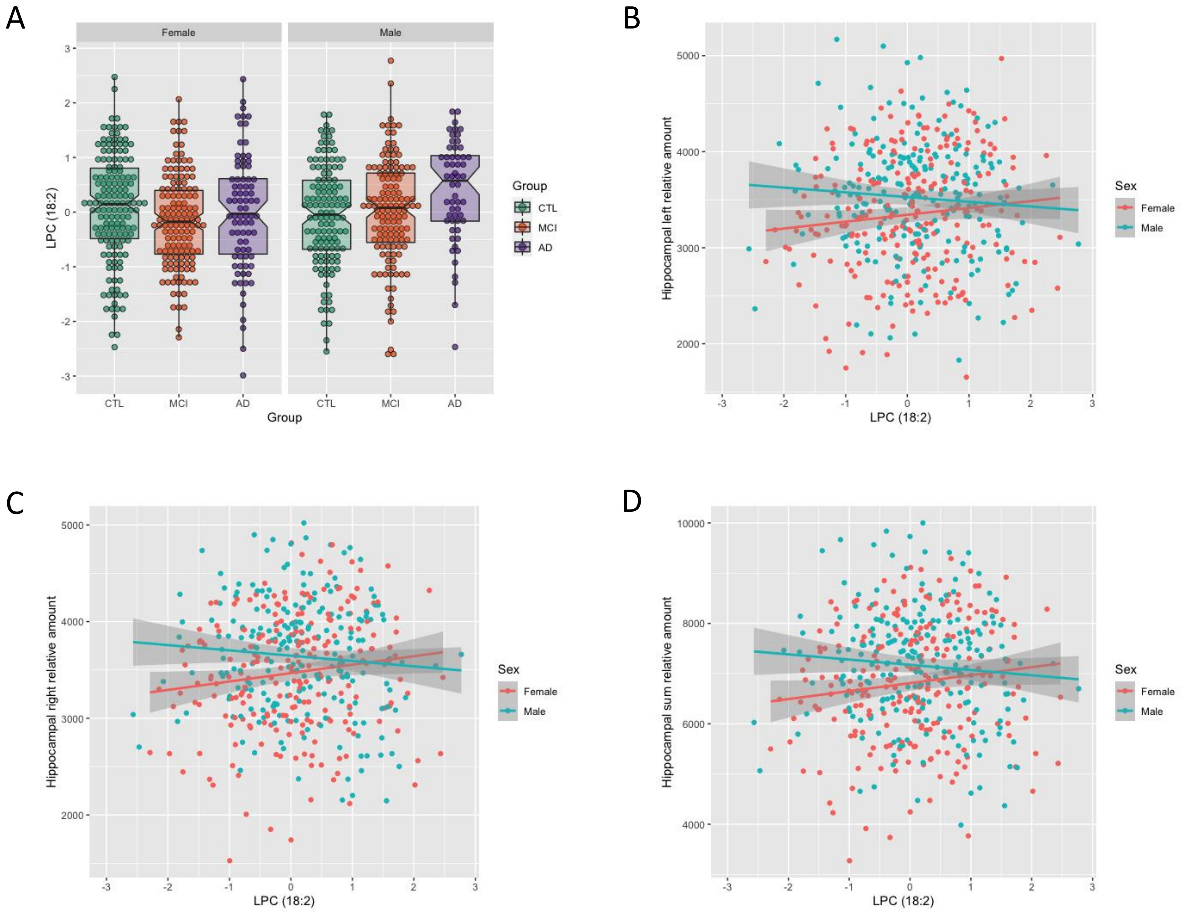


**Figure S9.** LPC (18:2) showed significant association in the male group with A) diagnosis (AD/CTL) (β = 0.69, 95% CI = 0.29 to 1.13, P = 1.1e-03), B) hippocampal left relative amount (β = -91.03, 95% CI = -170.63 to -11.43, P = 2.6e-02); C) hippocampal right relative amount (β = -98.73, 95% CI = -176.98 to -20.49, P = 1.5e-02) and D) hippocampal total relative amount (β = -189.75, 95% CI = -340.19 to -39.30, P = 1.42e-02).

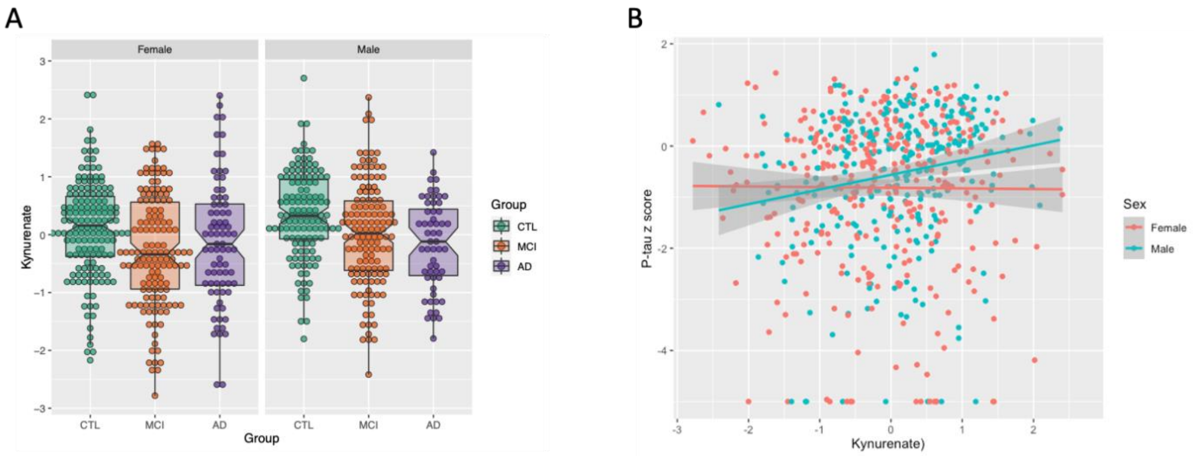


**Figure S10.** Kynurenate showed significant association in the male group with A) diagnosis (AD/CTL) (β = -1.04, 95% CI = -1.58 to -0.54, P = 7.6e-05) and B) p-tau z-score (β = 0.29, 95% CI = 0.11 to 0.47, P = 1.77e-03).

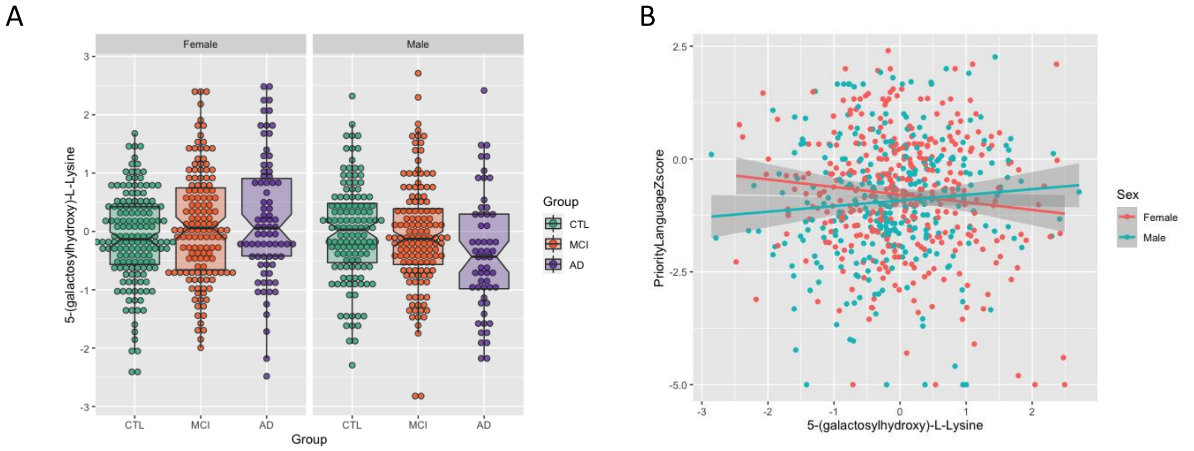


**Figure S11.** 5-(galactosylhydroxy)-L-Lysine showed significant association in the male group with A) diagnosis (AD/CTL) (β = -0.68, 95% CI = -1.12 to -0.26, P = 2.0e-03) and B) language z-score (β = 0.18, 95% CI = 0.0062 to 0.35, P = 4.3e-02).

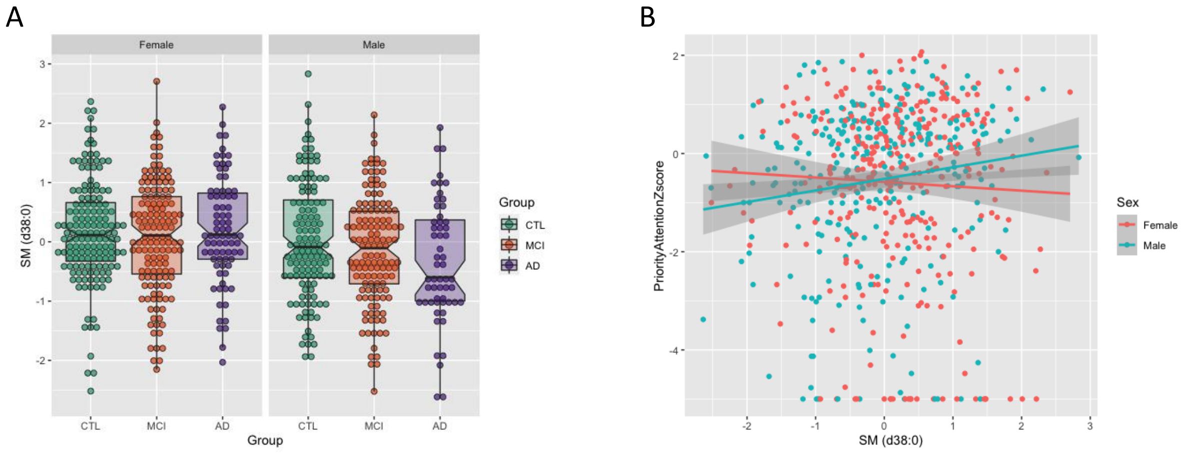


**Figure S12.** SM (d38:0) showed significant association in the male group with A) diagnosis (AD/CTL) (β = -0.47, 95% CI = -0.84 to -0.11, P = 1.2e-02) and B) attention z-score (β = 0.20, 95% CI = 0.016 to 0.39, P = 3.4e-02).

A

| | Reference + VMA + Tryptophan betaine | | Total | |
|---|---|---|---|---|
| Reference | <36.8% | >=36.8% | | |
| event | | | | $NRI_{event}$ |
| <36.8% | 7 | 6 | 13 | =(6-3)/75 |
| >=36.8% | 3 | 59 | 62 | =0.04 |
| New total | 10 | 65 | 75 | |
| Non-event | | | | $NRI_{non\text{-}event}$ |
| <36.8% | 103 | 4 | 107 | =(9-4)/129 |
| >=36.8% | 9 | 13 | 22 | =0.0388 |
| New total | 112 | 17 | 129 | |
| | | | | NRI = 0.0788 |

B

| | Reference + Kynurenate | | Total | |
|---|---|---|---|---|
| Reference | <34.7% | >=34.7% | | |
| event | | | | $NRI_{event}$ |
| <34.7% | 9 | 0 | 9 | =(0-0)/52 |
| >=34.7% | 0 | 43 | 43 | =0 |
| New total | 9 | 43 | 52 | |
| Non-event | | | | $NRI_{non\text{-}event}$ |
| <34.7% | 79 | 3 | 82 | =(3-3)/98 |
| >=34.7% | 3 | 13 | 16 | =0 |
| New total | 82 | 16 | 98 | |
| | | | | NRI = 0 |

C

| | Reference + VMA + Tryptophan betaine | | Total | |
|---|---|---|---|---|
| Reference | <35.9% | >=35.9% | | |
| event | | | | $NRI_{event}$ |
| <35.9% | 13 | 8 | 21 | =(8-2)/127 |
| >=35.9% | 2 | 104 | 106 | =0.0472 |
| New total | 15 | 112 | 127 | |
| Non-event | | | | $NRI_{non\text{-}event}$ |
| <35.9% | 183 | 8 | 191 | =(8-6)/227 |
| >=35.9% | 9 | 27 | 36 | =0.0044 |
| New total | 192 | 35 | 227 | |
| | | | | NRI = 0.0516 |

D

| | Reference + Kynurenate | | Total | |
|---|---|---|---|---|
| Reference | <35.9% | >=35.9% | | |
| event | | | | $NRI_{event}$ |
| <35.9% | 21 | 0 | 21 | =(0-1)/127 |
| >=35.9% | 1 | 105 | 106 | =-0.0079 |
| New total | 22 | 105 | 127 | |
| Non-event | | | | $NRI_{non\text{-}event}$ |
| <35.9% | 188 | 3 | 191 | =(3-3)/227 |
| >=35.9% | 3 | 33 | 36 | =0 |
| New total | 191 | 36 | 227 | |
| | | | | NRI = -0.0079 |

**Figure S13. Net reclassification index table for sex-specific biomarkers.** CSF biomarkers and covariates were used as baseline predictors in the reference models. The event, non-event, and total NRI were calculated for A) vanillylmandelate (VMA) and tryptophan betaine in the female sub-dataset; B) kynurenate in the male sub-dataset; C) VMA and tryptophan betaine in the full cohort; D) kynurenate in the full cohort.

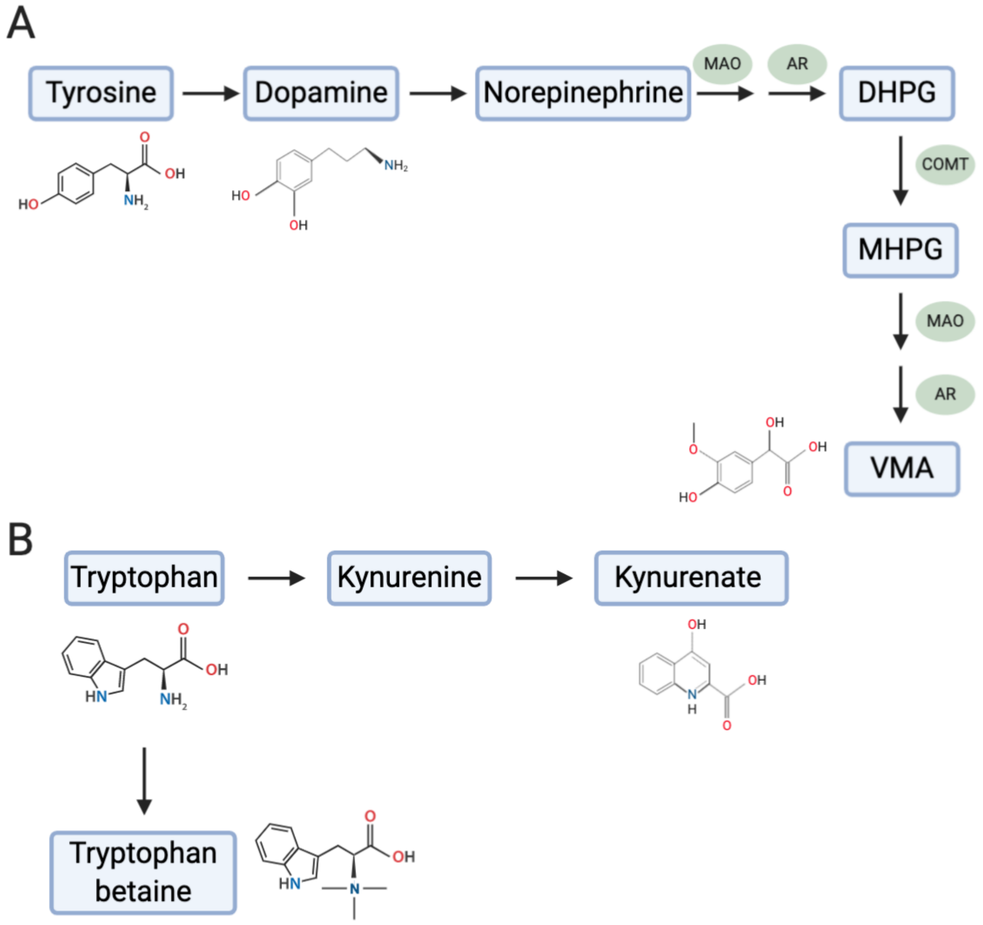


**Figure S14. VMA, tryptophan betaine, and kynurenate were generated through tyrosine and tryptophan pathways.** A) VMA is an end product of catecholamine metabolism in the tyrosine pathway metabolized by COMT, AR and MAO; B) kynurenate is produced from tryptophan in kynurenine pathway, while tryptophan betaine is mainly generated in plants and through gut microbiota.

VMA, vanillylmandelate; MAO, monoamine oxidase; COMT, catechol-O-methyl transferase; AR, aldehyde reductase; KAT, kynurenine aminotransferase. This figure is created with BioRender.com.

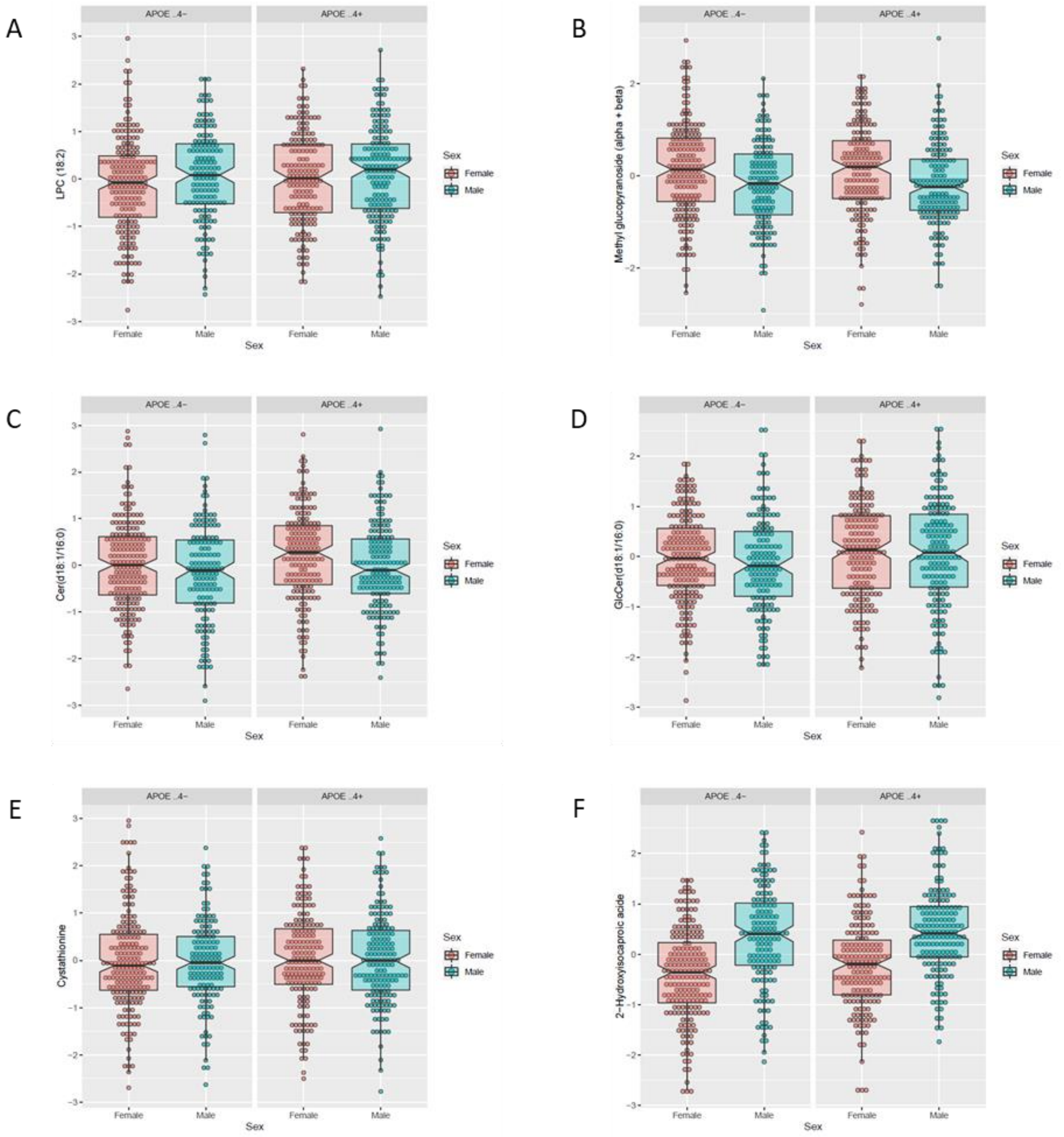


**Figure S15. Box plots of metabolites stratified by sex and *APOE* ε4 status.** A) LPC (18:2) and B) methyl glucopyranoside (alpha + beta) showed association with AD at nominal level in both male and *APOE* ε4 carrier strata; C) Cer(d18:1/16:0) and D) GlcCer(d18:1/16:0) showed association with visuoconstruction at nominal level in both male and *APOE* ε4 carrier strata; E) cystathionine showed association with attention at nominal level in both male and *APOE* ε4 carrier strata; F) 2-Hydroxyisocaproic acid showed association with immediate memory at nominal level in both female and *APOE* ε4 carrier strata.
